# Cost-effectiveness of CA125- and age-informed risk-based triage for ovarian cancer detection in primary care

**DOI:** 10.1101/2025.03.31.25324934

**Authors:** Runguo Wu, Kirsten D. Arendse, Tooba Hamdani, Fiona M. Walter, Emma J. Crosbie, Borislava Mihaylova, Garth Funston

## Abstract

**Background:** In England, current practice is cancer antigen 125 (CA125) testing with pelvic ultrasound scan (USS) if CA125 is ≥35 U/mL for triage of women with suspected ovarian cancer (OC) in primary care. However, OC risk varies with CA125 level and age. The Ovatools model predicts OC risk based on age and CA125 levels to support primary care triage.

**Method:** We evaluated five alternative triage pathways for suspected OC in primary care, using a decision model. Two CA125-USS sequential pathways used Ovatools risk: 1-3% (subsequent USS) and ≥3% (urgent referral), or age-adjusted CA125 thresholds equivalent to Ovatools risks. Three pathways involved concurrent CA125-USS testing, with referral if abnormal USS *or* one of the following: (1) Ovatools risk ≥3%, (2) CA125 above the equivalent age-adjusted threshold or (3) CA125 ≥35U/mL. Clinical and cost-effectiveness was compared against current practice for women over and under 50 years of age.

**Results:** All alternative pathways increased benefits at age ≥50 years, at additional cost. The incremental cost-effectiveness ratios for CA125-USS sequential pathways were below 30,000 British pounds, dropping below 20,000 British pounds if the Ovatools threshold for USS was increased to 1.2-1.4% risk.

**Discussion:** For women ≥50 years, the Ovatools and equivalent age-adjusted threshold sequential pathways are cost-effective compared to current practice.

## Background

In the UK, around 7,500 women are diagnosed with ovarian cancer (OC) every year [1]. There is no national screening programme for OC with large trials failing to demonstrate mortality benefit from screening [2, 3]. The majority of women are diagnosed after presenting in primary care with relevant symptoms [4], and most are not diagnosed until the disease is advanced when it is harder to treat [1]. The survival rate for OC varies significantly depending on the stage at diagnosis with 5-year survival rates of 95% and 16% for patients diagnosed at stage I and IV, respectively [5].

In the UK, OC diagnostic tests in primary care are limited to transvaginal/pelvic ultrasound scans (USS) and the serum biomarker Cancer Antigen 125 (CA125) [6]. The UK National Institute for Health and Care Excellence (NICE) guidelines for cancer detection (NG12) recommend women presenting in primary care with symptoms of possible OC be tested for CA125, followed by pelvic USS if CA125 ≥ 35 U/mL [7]. Although the 35 U/mL CA125 threshold performs well overall, the risk of OC, and other cancers, varies markedly depending on the individual’s CA125 level and age [8].

The Ovatools prediction models, developed and validated for use in primary care, use both CA125 levels and age to predict OC probability [8–10]. Ovatools estimate the probability of an invasive OC diagnosis within 12-months of CA125 testing in primary care. By providing a more individualised assessment of cancer risk, Ovatools could be used to select women for further cancer investigation in line with established national risk thresholds, and improve the diagnostic pathway for OC. Alternatively, age-adjusted CA125 thresholds, derived from the Ovatools risk models, could be used in place of the single CA125 threshold (currently used for all ages) to improve risk-based triage.

The current NICE recommendations for primary care OC detection were informed by a cost-utility decision analysis which considered combinations of the single cut-off of CA125 at 35 U/mL, USS and pelvic exam, and suggested that a CA125-USS sequential pathway was the most cost-effective strategy [11]. The decision model assumed a homogeneous target population and did not account for the interdependencies between age, cancer risk, diagnosis and outcomes and, therefore, cannot inform an assessment of the value of Ovatools in clinical practice.

In this study, we present a new decision analytic model informed by estimates of cancer diagnosis, outcomes, survival, and costs in a large UK primary care population of women tested using CA125 and/or USS. We used this model to evaluate the cost-effectiveness of new Ovatools-based OC detection pathways in primary care with the ultimate aim of improving outcomes from OC.

## Methods

The study focused on assessing pathways to detect invasive OC, excluding borderline tumours (see **Supplementary Table S1** for ICD-10 codes), in primary care.

### Pathways for OC diagnosis in primary care

We considered six pathways for OC diagnosis among women presenting in primary care with symptoms of possible OC. Firstly, we examined the NICE recommended pathway (Pathway 1) of CA125 test, followed by pelvic USS if CA125 ≥ 35 U/mL (**Figure 1A**).

**Figure 1:**
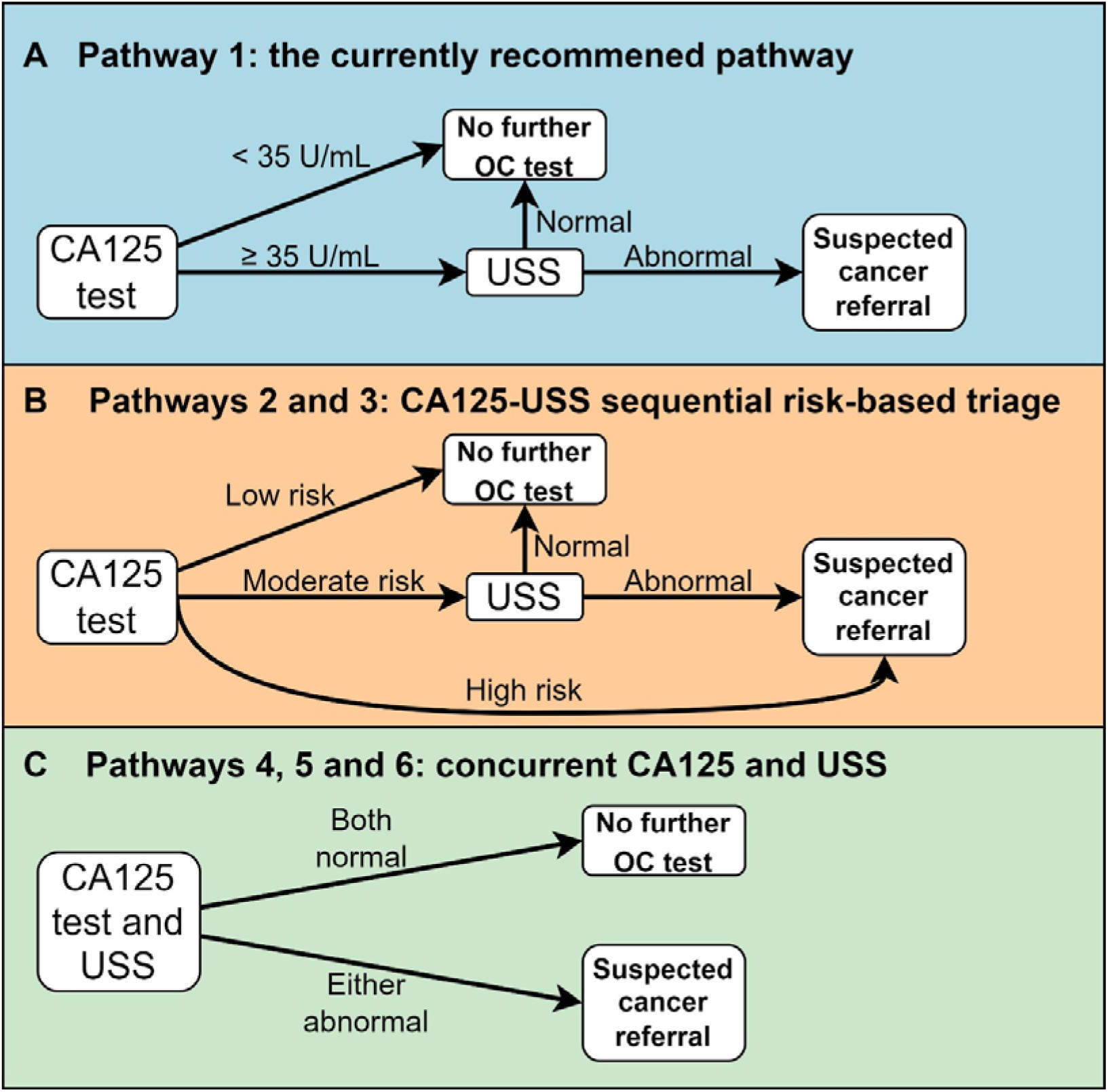
Ovarian cancer detection pathways in primary care. **Pathway 1** is the currently recommended CA125-USS sequential pathway in England. In the risk-based triage pathways, we used Ovatools risk <1%, 1-3% and ≥3% in **Pathway 2**, or their equivalent age-adjusted CA125 thresholds in **Pathway 3**, to indicate low, moderate and high risks, respectively. In pathways with concurrent CA125 and USS, the abnormal CA125 threshold is Ovatools risk ≥3% (**Pathway 4)**, its equivalent age-adjusted CA125 threshold (**Pathway 5)**, or CA125 ≥35 U/mL (**Pathway 6)**. CA125, cancer antigen 125; OC, ovarian cancer; USS, pelvic/transvaginal ultrasound scan.

Secondly, two further CA125-USS sequential pathways using Ovatools-informed [10] risk-based triage or equivalent age-adjusted CA125 thresholds were studied (**Figure 1B**). In Pathway 2, patients first undergo a CA125 test and Ovatools risk is calculated. If the Ovatools invasive OC probability is <1% patients are not investigated further for OC; if it is between 1% and 2.9% (moderate risk) they are followed up with pelvic USS; and if ≥3% (high risk) they are directly referred on an urgent suspected OC pathway for further assessment. The 1% cancer risk threshold has been used by NICE when selecting symptomatic patients for primary care investigations (such as chest X-ray in patients with symptoms of possible lung cancer) and the 3% threshold aligns with the level recommended for urgent suspected cancer referrals in symptomatic patients [7]. Pathway 3 mimics Pathway 2 but instead uses age-adjusted CA125 thresholds with OC detection accuracy matching the corresponding Ovatools probabilities of ∼1% and ∼3% (see **Supplementary Method 1** for details of the age group specific thresholds and their accuracy) [10].

Thirdly, three further CA125-USS concurrent pathways were studied, where patients undergo both CA125 testing and a pelvic USS in primary care (**Figure 1C**). In Pathway 4, patients are referred if either the Ovatools invasive OC probability is ≥3% or OC is suspected on USS. Pathway 5 uses age-adjusted CA125 threshold matching Ovatools probability of 3% and Pathway 6 uses CA125≥35 U/mL threshold in place of the Ovatools probability.

### Study populations

This study included women with records indicating they underwent a CA125 test and/or a pelvic/transvaginal USS between April 1st, 2013, and December 31st, 2017, in Clinical Practice Research Datalink (CPRD) Aurum. The index date for the study was the earliest valid CA125 test or USS record within the study period. CPRD Aurum is a database of de-identified coded primary care records, which contains routinely-collected data from 40 million patients from 1500 UK general practices which use the EMIS Web® electronic patient record system software [12]. Data from CPRD Aurum was linked at the patient level to Hospital Episode Statistics (HES), death registration data and National Cancer Registration and Analysis Service (NCRAS) data [13]. CA125 records were obtained from CPRD Aurum, and USS records were obtained from CPRD Aurum and linked HES Diagnostic Imaging Data (HES DID).

Women with a previous USS or CA125 test within 365 days before the index date or with a record for OC (including borderline ovarian tumour) at any time before the index dates were excluded. Women with a USS record but neither a CA125 record nor ovarian related symptoms were also excluded as the indication for USS is not always recorded within CPRD. Patients were followed until March 31st, 2021 [14]. As in previous studies, we assumed all patients with cancer at the time of CA125 testing or USS were diagnosed within 1 year [15].

The base-case cost-effectiveness analysis used a narrower target population of CA125 tested women, while certain model parameters were estimated using the broader study population.

### The decision analytic model

The decision analytic model consists of a decision tree to represent the diagnostic pathways and costs in primary care, and a Markov model to capture long-term survival, quality of life (QoL) and inpatient care costs. The structure aligns with the model that informed NICE recommendations [11]. The decision tree estimates patients’ probabilities of the following outcomes after assessment in primary care, namely, (1) invasive OC detected and referred to secondary care (true positive), (2) invasive OC undetected and patient reassured (false negative), (3) no invasive OC and reassured (true negative), and (4) no invasive OC but referred to secondary care (false positive). To parametrise the decision tree, we used the accuracy data for CA125 (with a 35 U/mL threshold) and Ovatools, stratified by age (<50 years and ≥50 years), as the accuracy changes significantly with age (**Table 1**) [10]. We applied the same USS accuracy metrics as used in the study informing NICE and the economic evaluation of OC pathways in UK secondary care [11, 16]. Disease incidence and stage were estimated based on patient characteristics from the study population using logistic regression models (**Supplementary Method 2**).

**Table 1:** Parameters in the decision tree model of primary care pathway.

| Parameters | Value (95% CI) | Source |
| --- | --- | --- |
| **Test accuracy** |  |  |
| CA125 35 U/mL sensitivity, age 18-49 years | 75.3 (70.0 – 80.0) | a |
| CA125 35 U/mL specificity, age 18-49 years | 92.5 (92.3 – 92.6) | a |
| Ovatools 1% sensitivity, age 18-49 years | 61.9 (56.1 – 67.4) | a |
| Ovatools 1% specificity, age 18-49 years | 96.9 (96.8 – 97.0) | a |
| Ovatools 3% sensitivity, age 18-49 years | 45.2 (39.4 – 51.0) | a |
| Ovatools 3% specificity, age 18-49 years | 99.4 (99.3 – 99.4) | a |
| CA125 35 U/mL sensitivity, age ≥50 years | 86.5 (84.8 – 88.0) | a |
| CA125 35 U/mL specificity, age ≥50 years | 94.3 (94.2 – 94.4) | a |
| Ovatools 1% sensitivity, age ≥50 years | 91.1 (89.7 – 92.4) | a |
| Ovatools 1% specificity, age ≥50 years | 89.1 (88.9 – 89.2) | a |
| Ovatools 3% sensitivity, age ≥50 years | 83.1 (81.3 – 84.8) | a |
| Ovatools 3% specificity, age ≥50 years | 96.5 (96.4 – 96.6) | a |
| Ultrasound sensitivity, all ages (%) | 85 (83 – 87) | [11]b |
| Ultrasound specificity, all ages (%) | 83 (81 – 85) | [11]b |
| **Pathway effect on OC** |  |  |
| Proportional risk reduction in late-stage OC incidence if detected earlier than usual care | 0.836 (0.737 – 0.950) | [18]c |
| **Primary care costs** |  |  |
| CA125 test cost | £10 | [19] |
| Cost of nurse time for blood test (10 min) | £9 | [20] |
| Average USS test cost | £204 | [21]d |
| Cost of GP face-to-face consultation (10 min) | £40 | [20] |
| Cost of GP telephone consultation (5 min) | £20 | [20] |
| Cost of outpatient consultation | £181 | [21] |
| **Parameters related to benign disease** |  |  |
| Cost of benign disease surgery | £3,994 | [21] |
| Surgery rate for benign disease after referral | 0.75 | e |
| QoL disutility in the year of surgery | 0.04 (0.01 – 0.06) | [22] |
| Annual QoL utility gain following surgery after year of surgery | 0.008 (-0.005 – 0.021) | [23] |

a. Informed by a parallel study using CPRD data [10]; b. Data used in the health economic evaluation informing the NICE clinical guideline CG122[11, 24]; c. data for symptomatic women; d. activity-weighted average cost across outpatient services and directly accessed diagnostic services; e. Among referred non-cancer patients with positive USS for OC detection. Informed by ROCKeTS data [25]. CA125, cancer antigen 125; CI, confidence interval; USS, ultrasound scan; OC, ovarian cancer; GP, general practitioner.

We estimated the effect of higher cancer detection rates in primary care by modelling the cancer stage shift [17]. We applied it to patients who were diagnosed at the late stage of cancer if they were in the false negative group in the current pathway but in the true positive group in the new pathway. We assumed that a fraction of additionally detected ‘previously late stage’ cancers would ‘shift’ to an early stage at diagnosis in the new pathway. This fraction was estimated using the relative risk ratio of late-stage diagnosis incidence between screen detected cases and clinically detected cases among symptomatic women in the UKCTOCS study [18]. We assumed late-to-early shifted cancer cases to have the same risk of cancer death as that of an early-stage cancer diagnosis. (**Supplementary Method 3**).

The Markov model includes five states: three entry states – no cancer, early-stage cancer, and late-stage cancer – and two absorbing states – death caused by cancer (cancer death) and death not caused by cancer (non-cancer death) (**Supplementary Figure S1**). The model uses annual cycles. The transition probabilities from the cancer states to cancer death were estimated using flexible parametric survival models, which modelled time from cancer diagnosis to cancer death using data from the study population. As a high proportion of patients with raised CA125 have other cancers [8], the survival models were estimated separately for OC and cancers including lower gastrointestinal (GI) cancer, uterine cancer, lung cancer and pancreatic cancer, and other cancer types were modelled as a whole. Covariates included age, cancer stage at diagnosis, ethnicity, and socioeconomic deprivation.

Missing cancer stage data was imputed using multiple imputation prior to model estimation (**Supplementary Method 4**). No detailed cancer progression was modelled, but the models of cancer death risk subsume cancer progressions. The estimated survival models were used to predict cancer death for cancer patients in the first eight years after diagnosis in the Markov model. The national statistics of mortality rates by age, sex and cause (cancer and non-cancer) were used to predict separately all non-cancer death and cancer death beyond eight years. The models showed good performance in validation on internal and external data (**Supplementary Method 5**).

### Cost estimation

Three categories of costs, in 2022 British pounds (£), were included in the analyses. Firstly, we considered the costs for services and diagnostic tests in primary care pathways. These include the costs for the general practitioner (GP) time, nurse time, the CA125 test, and the USS (see **Table 1** for cost details and sources). We assumed that each patient received an initial face-to-face GP consultation, followed by a CA125 test (and USS in concurrent pathways). In sequential pathways, if patients’ CA125 test results indicated an USS, they received a GP telephone follow-up to inform them of the result and arrange an USS appointment, and if the USS was abnormal, they received a further GP consultation before being referred to secondary care. Test results indicating no need for further action in the pathway did not lead to additional GP consultations. In cases where cancer existed but was not detected in the initial diagnostic pathway, we assumed the diagnostic process in primary care was repeated once.

Secondly, we estimated the long-term hospital inpatient care costs from hospital records, including cancer treatment, using the Hospital Episode Statistics (HES) Health Resource Group (HRG) data. HRG codes were mapped to NHS reference costs inflated to 2022 prices [26]. Following methods used previously [27, 28], separate Two-Part Generalised Linear Models were fitted for patients with a certain cancer type as those in the survival models, and patients without cancer diagnosed. Duration since the first diagnosis was defined as a categorical variable with annual levels from the year of diagnosis to four or more years after diagnosis. Cancer stage at diagnosis, current age, ethnicity and socioeconomic deprivation were included as covariates. These cost models were integrated into the Markov model for prediction of inpatient care costs since diagnosis.

Finally, we added hospital costs for referrals without an eventual cancer diagnosis, because the above inpatient care cost models do not effectively capture hospital cost for those referred to hospital but not ultimately diagnosed with cancer. We separately added costs for patients who did and did not undergo surgery for benign diseases. We used data from the ROCkeTS study to determine the surgery rate among patients referred from primary care who were not diagnosed with any cancer (**Table 1**), and applied the activity-weighted cost of “Treatment with interventions for Non-Malignant Gynaecological Disorders” in the NHS reference costs [26]. For non-surgery cases, we assumed all patients received an outpatient consultation, a CA125 test and a USS (see **Supplementary Method 6** for further details about cost estimation).

### Quality of life

We estimated QoL associated with patient characteristics and disease histories using a linear regression model of EuroQoL-5 Dimension (EQ-5D) utility using data from UK Biobank [29]. The EQ-5D utility generally ranges from around −0.5 for the worst health state to 1 for full health where 0 is equivalent to death [30]. Histories of different cancers were defined as categorical variables with levels of no such cancer, cancer diagnosed within one year, and cancer diagnosed more than one year previously.

Ethnicity, Townsend score quintiles and age were included as covariates. The QoL model was integrated into the Markov model for prediction of QoL over lifetime and estimation of quality adjusted life years (QALYs). See **Supplementary Method 7** for further details. As UK Biobank does not provide data for cancer stage, a QoL decrement of 0.046 derived from literature was used to adjust QoL for any late-stage cancer [31]. For patients referred to secondary care without a cancer diagnosis, if they received surgery, we assumed a reduction of 0.04 in QoL in the year of surgery [22], and an improvement of 0.008 in annual QoL thereafter [23] (**Table 1**).

### Cost-effectiveness analysis

Base-case analysis focused on women with CA125 records to inform the real-world cost effectiveness of implementing Ovatools thresholds in place of the current CA125 cut-off, while we also conducted analysis on the wider population including also symptomatic women with USS but without CA125 testing. We conducted cost-effectiveness analyses from the perspective of the UK health services. We used the decision model to simulate diagnostic outcomes, remaining life years (LYs), QALYs and costs for the study populations over a lifetime (or until 110 years of age) assuming they followed a particular primary care pathway. We applied a 3.5% annual discount rate to costs and QALYs.

We used two approaches to compare the results of different pathways. First, we compared the NICE recommended Current Threshold Sequential pathway with each alternative pathway to derive QALYs gained and additional costs and identified dominated alternatives. Second, we calculated incremental cost-effectiveness ratios (ICERs) based on comparisons of moving to increasingly effective and costly alternative pathways, excluding the dominated alternatives [32]. We did not directly compare the Ovatools pathways with their corresponding age-adjusted Threshold pathways, as the differences between them were minor due to the age-group specific thresholds being aligned with Ovatools’ risk-based accuracy.

Given that the sensitivity of Ovatools at the 1% risk threshold is lower than that of CA125 at the 35 U/mL threshold for individuals aged < 50 years but higher for those aged ≥ 50, we analysed the data separately for these two age groups.

### Sensitivity and scenario analyses

Parameter uncertainty was assessed using probabilistic sensitivity analysis based on 1000 model parameter sets generated using bootstrap methods or Monte Carlos simulations (See **Supplementary Method 8** for details). With the probabilistic data, the cost-effectiveness acceptability curve was produced by comparing the net benefits among selected pathways at a range of cost-effectiveness thresholds.

In one-way sensitivity or scenario analyses, first, we applied the lower and higher bounds of the 95% confidence interval of the relative risk ratio of late-stage diagnosis (**Table 1**) to estimate the stage shift effect, respectively. Second, we increased or decreased the sensitivity and specificity of USS by 5%, respectively. Third, we used the cost for USS in directly accessed diagnostic services or the cost for USS in outpatient care, while the base case used the weighted combination of the two. Fourth, we excluded the short-term QoL reduction or the long-term QoL improvement due to benign gynaecological surgery one at a time. Fifth, we increased or decreased the surgery rate for referred non-cancer cases by 15%. Sixth, we used decrement of 0.06 informed from NHS Cancer Quality of Life Survey [33] or zero decrement, to adjust QoL utility for late-stage cancer. Seventh, we included the pathway’s effects on lower GI, uterine, lung and pancreatic cancers (See **Supplementary Method 9** for details). Eighth, we used age- and cancer stage-specific CA125 and Ovatools accuracy data to replace the age-specific accuracy data in the decision tree (**Supplementary Method 10**). Ninth, we extended the use of estimated cancer death survival models from first eight to 15 years. Tenth, we used 1.5% discount rates for both outcomes and costs. The details of sensitivity analyses are summarised in **Supplementary Table S2.**

Finally, we calculated the cost-effectiveness of the *Ovatools Sequential pathway* compared with *the Current Threshold Sequential pathway* across different Ovatools risk thresholds from 0.6% to 4% (**Supplementary Method 11**).

### Patient and Public Involvement

We held a patient and public involvement (PPI) workshop, including PPI members with experience of CA125 testing and ovarian cancer, and shared preliminary findings within this group. PPI members’ perspectives on key results and potential implications for patients and the public were gathered to inform manuscript preparation.

### Statistic software

All the analyses were performed using R 4.2.3.

## Results

### Summary of characteristics

The base case analysis included 276,827 women who underwent CA125 testing, with an average age of 54.6 years. 89.9%, 5.7% and 3.4% were White/White British, Asian/Asian British and Black/Black British ethnicity, respectively. 1,964 (0.71%) were diagnosed with invasive OC within 1 year of CA125 testing. Of those with stage recorded at diagnosis, 68.5% had late-stage OC (III-IV). 0.48%, 0.25%, 0.21%, 0.16% and 1% were diagnosed with lower GI cancer, uterine cancer, lung cancer, pancreatic cancer and other cancers, respectively, within 1 year of CA125 testing (**Table 2).** The population characteristics of women with CA125 or USS records is presented in **Supplementary Table S3**.

**Table 2:** Characteristics of the base-case analysis population.

|  | Mean (SD) or n (%) |
| --- | --- |
| N | 276,827 |
| Age | 54.6 (15.8) |
| Ethnicity |  |
| White | 248,957 (89.9) |
| Asian | 15,693 (5.7) |
| Black | 9,379 (3.4) |
| Mixed or others | 2,798 (1.0) |
| Townsend Score |  |
| Q1 (least deprived) | 67,437 (24.4) |
| Q2 | 60,760 (21.9) |
| Q3 | 53,900 (19.5) |
| Q4 | 46,959 (17.0) |
| Q5 | 47,771 (17.3) |
| Incident cancer diagnosed within 1 year |  |
| Invasive Ovarian cancer | 1,964 (0.71) |
| Stage 1/2 | 444 (22.6) |
| Stage 3/4 | 965 (49.1) |
| Stage missing | 555 (28.3) |
| Lower GI cancer | 1,325 (0.48) |
| Uterine cancer | 691 (0.25) |
| Lung cancer | 593 (0.21) |
| Pancreatic cancer | 441 (0.16) |
| Other cancers | 2,820 (1.0) |
| Duration of follow up (years) | 5.40 (1.58) |

### Pathway diagnostic outcomes

Among CA125 tested women aged < 50 years, 0.2% were diagnosed with invasive OC within 1 year, and of those with recorded stage, 46% had late-stage disease. The model predicted that the currently recommended pathway (Pathway 1), the two new CA125-USS sequential pathways using Ovatools and age-adjusted thresholds, respectively (Pathways 2 and 3), and the three CA125-USS concurrent pathways using Ovatools, age-adjusted and current CA125 thresholds, respectively (Pathways 4, 5 and 6) detected 64%, 59%, 60%, 92%, 92% and 96% of invasive OC, and referred 1.4%, 1.1%, 1.2%, 18%, 18% and 23% of patients to further secondary care assessment, respectively.

Among CA125 tested women aged ≥50 years, 1.05% had invasive OC diagnosed within 1 year, and of those with recorded stage, 72% had late-stage disease. Pathways 1, 2, 3, 4, 5 and 6 were predicted to detect 74%, 90%, 90%, 97%, 97% and 98% of invasive OC, and referred 1.7%, 5.7%, 5.8%, 21%, 21% and 23% of patients to further secondary care assessment, respectively (**Table 3**).

**Table 3:** Outcomes, costs and cost-effectiveness, by primary care pathway.

| Pathway | Invasive OC detection rate (%) | Referral rate, among all patients (%) | QALYs gained per 1000* | Cost difference per 1000 (£)* | ICER (£/QALY gained)** |
| --- | --- | --- | --- | --- | --- |
| **Age ≤ 49 years:** N = 112,081; 228 (0.2%) invasive OC with 46% late stage |  |  |  |  |  |
| **Pathway 1 (current practice)** | 64 | 1.4 |  |  |  |
| Pathway 2 | 59 | 1.1 | -0.97 | -33,354 | [-34,350] |
| Pathway 3 | 60 | 1.2 | -0.95 | -33,455 | [-35,348] |
| Pathway 4 | 92 | 18 | 0.3 | 258,083 | ED |
| Pathway 5 | 92 | 18 | 0.33 | 259,095 | ED |
| Pathway 6 | 96 | 23 | 2.44 | 334,595 | 137,123 |
| **Age ≥ 50 years:** N = 164,746; 1736 (1.05%) invasive OC with 72% late-stage |  |  |  |  |  |
| **Pathway 1 (current practice)** | 74 | 1.7 |  |  |  |
| Pathway 2 | 90 | 5.7 | 1.48 | 34,894 | 23,610 |
| Pathway 3 | 90 | 5.8 | 1.53 | 39,327 | 25,712 |
| Pathway 4 | 97 | 21 | 1.86 | 283,225 | ED |
| Pathway 5 | 97 | 21 | 1.86 | 283,223 | ED |
| Pathway 6 | 98 | 23 | 2.23 | 304,856 | 358,960 |

\*Compared with Pathway 1; QALYs and cost were discounted. \*\*Compared with the non-dominated less costly alternative (Ovatoools and its equivalent age-adjusted threshold pathways were not compared with each other). ICERs in square brackets indicate £ saved per 1 less QALY, due to reduction in benefit and cost saving compared to Pathway 1.
CA125, cancer antigen 125; OC, ovarian cancer; QALY, Quality-adjusted life year; ICER, incremental cost-effectiveness ratio; USS, ultrasound

### Cost-effectiveness

Per 1000 CA125 tested women aged < 50 years, compared with Pathway 1, the two new CA125-USS sequential pathways (Pathways 2 and 3) resulted in 0.97 and 0.95 fewer QALYs and cost savings of £33,354 and £33,455, respectively. Conversely, the three CA125-USS concurrent pathways (Pathways 4, 5 and 6) resulted in 0.3, 0.33 and 2.44 extra QALYs, with additional costs of £258,083, £259,095 and £334,595, respectively. Following the exclusion of the dominated alternatives, compared to Pathway 1, Pathway 6 resulted in more QALYs at £137,123 per QALY gained (**Table 3**).

Per 1000 CA125 tested women aged ≥ 50 years, compared with Pathway 1, Pathways 2, 3, 4, 5 and 6 resulted in 1.48, 1.53, 1.86, 1.86 and 2.23 more QALYs, at additional costs of £34,894, £39,327, £283,225, £283,223 and £304,856, respectively.

The additional costs in the two new sequential pathways (Pathways 2 and 3) were driven by secondary care costs due to increased referrals in false positive cases. In contrast, the additional costs in the concurrent pathways (Pathways 4, 5 and 6) were driven by the primary care diagnostic costs, where all women underwent USS (**Supplementary Table S4**). Following the exclusion of the dominated alternatives, compared to Pathway 1, the additional QALYs with Pathways 2 and 3 costed £23,610 and £25,712 per QALY gained, and compared to Pathway 2, the additional QALYs with Pathway 6 costed £358,960 per QALY gained (**Table 3**).

The outcomes in the broader population of women with CA125 or USS show similar patterns (**Supplementary Table S5 & S6**).

The cost-effectiveness acceptability curves indicate that at £30,000 per QALY gained threshold, among women aged ≥ 50 years, Pathways 2 and 3 had higher probability of being cost-effective compared with the other pathways including the currently recommended pathway (**Figure 2A, Supplementary Figure S2A, Supplementary Figure S3**).

**Figure 2:**
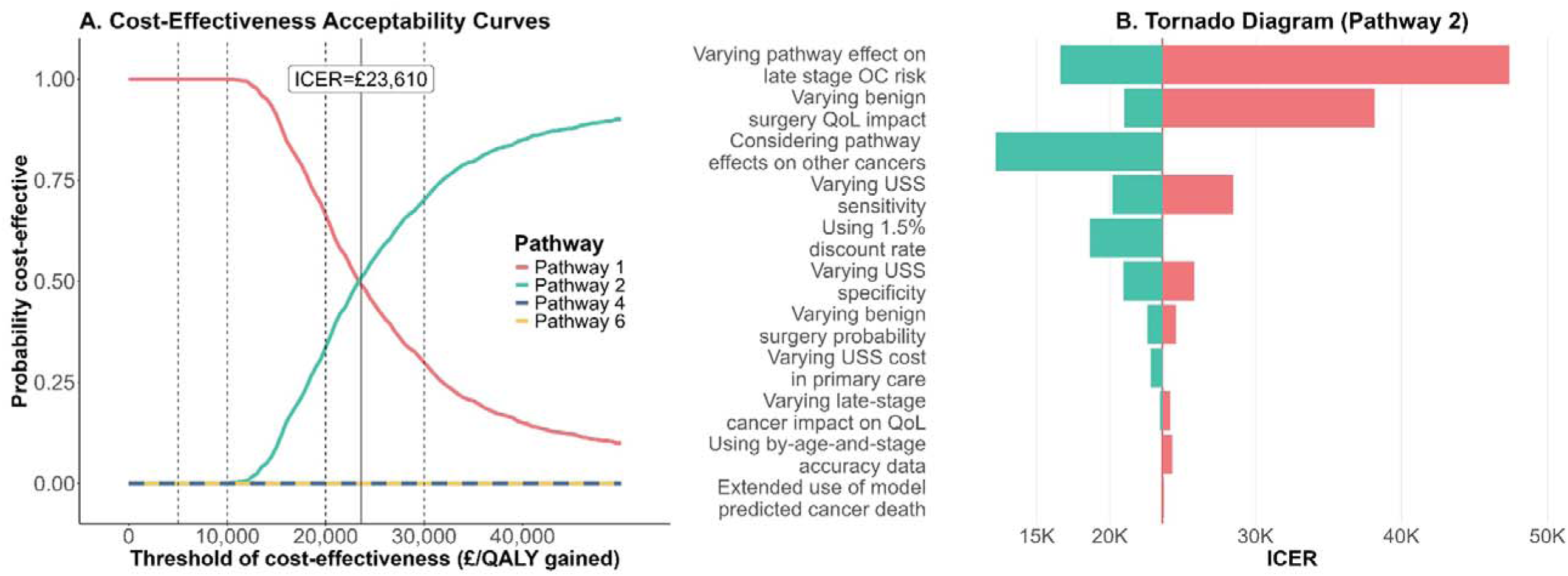
Probabilistic outcome and sensitivity analyses for CA125 tested women aged ≥ 50 years. Panel B: See supplementary Table S2 for the base case values and ranges used in the sensitivity or scenario analyses.

### Sensitivity and scenario analyses

In view of the promising findings for Pathways 2 and 3 among women aged ≥ 50 years, the one-way sensitivity and scenario analyses focused on Pathway 2; these results are generalizable to Pathway 3. The Tornado diagram shows that the ICER significantly increased with smaller reduction in late-stage OC risk due to earlier detection and exclusion of long-term QoL improvement associated with surgery for benign diseases. It also moderately increased with an increase in USS sensitivity.

Conversely, the ICER decreased to below £20,000 with greater reduction in late-stage OC risk due to earlier detection, with inclusion of the effects on other cancers, or with use of 1.5% discount rate (**Figure 2B, Supplementary Figure S2B & Supplementary Table S2**).

In a scenario analysis, increasing the moderate-risk threshold from 1% to 1.2% – 1.4% in Pathway 2 resulted in lower ICER to below £20,000. Further increases of the moderate-risk threshold up to 2% resulted in even lower ICER reaching negative values and indicating that benefits can be achieved while costs reduced. Changing the high-risk threshold has a minor impact on the ICER (**Figure 3 and Supplementary Table S7 & S8**). Similar observations were noted for Pathway 3 adjusting the age-group specific CA125 thresholds to correspond to respective Ovatools risk levels (**Supplementary Table S9**).

**Figure 3:**
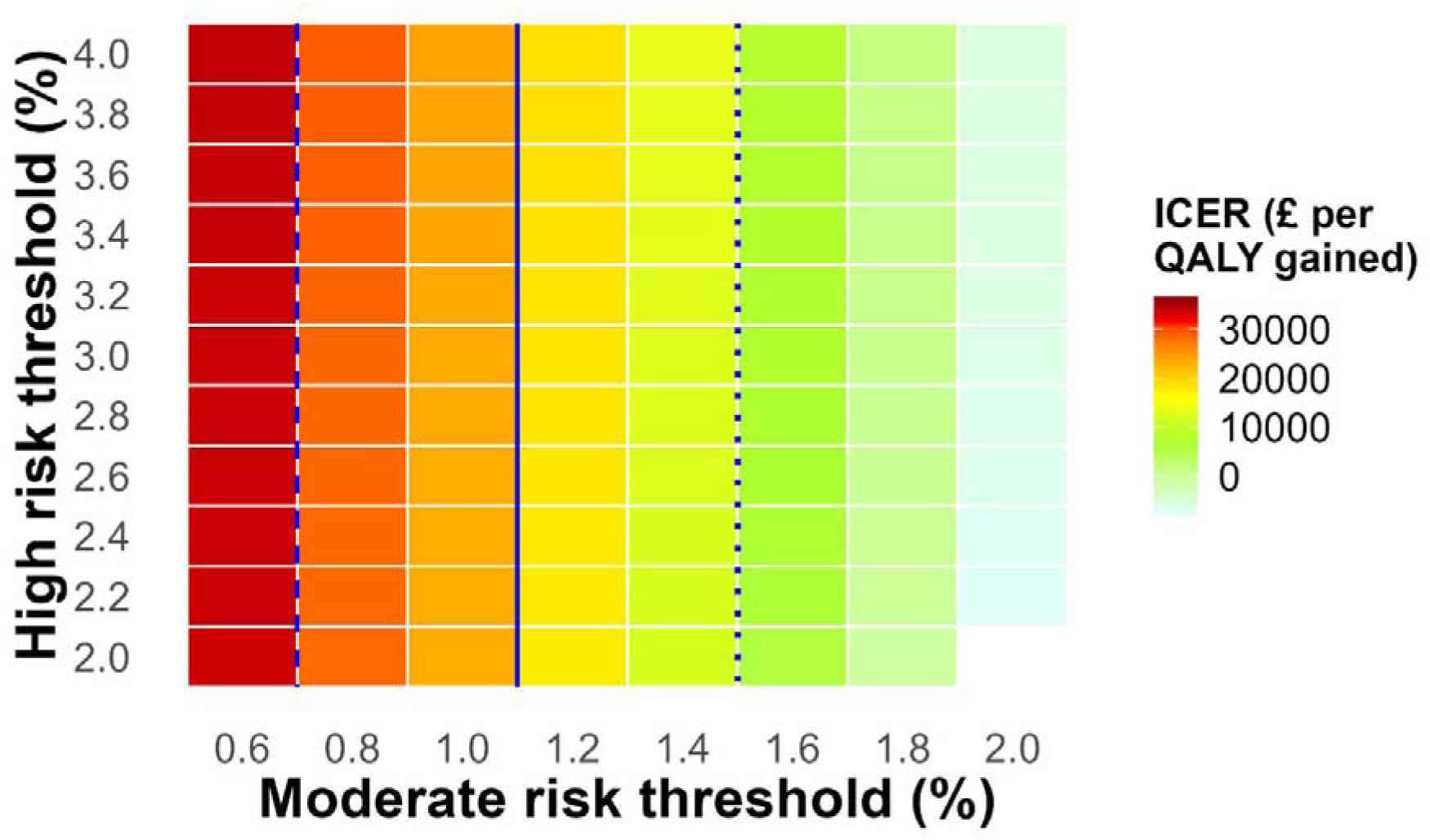
The heatmap of ICERs of the Pathway 2 compared with Pathway 1 with different risk thresholds for moderate and high risks, among CA125 tested women aged ≥ 50 years. The dotted, solid and dashed lines indicating the ICER thresholds of £10,000, £20,000 and £30,000 per QALY gained. ICER, incremental cost-effectiveness ratio; QALY, quality-adjusted life year.

## Discussion

We assessed the cost-effectiveness of alternative OC diagnostic pathways in real-world OC-suspected UK primary care population, based on OC risk estimated by CA125 levels and age using the Ovatools models or equivalent Ovatools-informed age-adjusted CA125 thresholds. Our findings indicate that, for women aged over 50 years, the modified CA125-USS sequential pathways are cost-effective at willingness-to-pay of £30,000 per QALY gained, using Ovatools probabilities of 1% and 3% as the thresholds, or its equivalent age-adjusted CA125 thresholds, for subsequent USS and urgent cancer referral to gynaecology, respectively. A small increase of the threshold for subsequent USS to 1.2% – 1.4% reduced ICER to below £20,000. For women under 50 years, only pathways with USS concurrent to CA125 improve OC detection but these were not cost-effective.

While numerous studies and a few cost-effectiveness analyses exist for OC screening and treatment [16, 19, 34–37], data on OC detection in primary care, particularly regarding cost-effectiveness, is limited. The most relevant evidence is from the study underpinning NICE CG122 [11] but this study is now over a decade old and lacks relevance for evaluating new pathways in clinical practice. Our study offers a more robust methodological framework for evaluating OC primary care diagnostic pathways, with the model code publicly available online. The analyses are grounded in representative UK primary care population data. Furthermore, the model builds upon the earlier studies [11, 16] guided by primary and secondary health care experts, ensuring clinical relevance. The CA125 and Ovatools accuracy data were provided by a parallel population-based cohort study using a similar population [10]. Parameters for predicting long-term mortalities, inpatient care costs and QoL, were derived using CPRD and its linked data, national statistics and UK Biobank, allowing for key patient characteristics, with cancer mortality prediction models validated both internally and externally. Through extensive sensitivity and scenario analyses, this study provides valuable cost-effectiveness data to inform decision makers considering the adoption of new clinical pathways for OC diagnosis in primary care in the UK and potentially other countries. It is noted that cost-effectiveness is improved when we considered the impact of pathways on lower GI, uterine, lung and pancreatic cancers, as evidence suggests that, in addition to OC, CA125 levels are often raised in women with a range of cancers, with the four included cancer types being particularly associated with CA125 elevations [8]. To our knowledge, this is the first attempt to quantify the broader impact of OC diagnostic pathways on other cancers.

Translating improvements in cancer detection to survival benefits is challenging. Most pathways have not yet been implemented in clinical practice, no clinical trials have evaluated other OC tests in primary care and no previous studies have examined the potential survival benefits from more timely OC detection within primary care. Instead, we drew on the best available data to inform our modelling. We translated the differences in OC detection across pathways into changes in late-stage diagnosis incidence, using screening data among symptomatic women in UKCTOCS trial [18], and linked the differences to changes in cancer mortality risk using survival models based on routine data. The major limitation of this method comes from the two assumptions. First, we assumed that symptomatic women in the UKCTOCS trial could represent OC-suspected patients in primary care, which may not be the case.

Secondly, we assumed the additional true positive cases in the new pathway to have the same late-stage OC risk reduction rate as that of the screen detected cases compared with the clinically detected cases. However, without direct evidence – which can take many years to obtain following trials or clinical implementation – this is the best approach to inform the cost-effectiveness of alternative pathways, in order to guide interventions aimed at improving OC detection in primary care in a timely manner.

Our sensitivity analysis shows that varying the risk reduction of late-stage OC impacts the ICER, but, notably, the 16% risk reduction may represent a conservative estimate, as another modelling study suggests a 71% reduction in late-stage OC due to early detection by screening [38]. It is known that the UKCTOCS randomised trial reported a significant reduction in late-stage OC incidence due to earlier detection by screening [18, 39], which, however, was not translated into a significant reduction in mortality [2, 40]. PLCO, another large randomised trial, also reported similar results [3]. However, this does not negate the link between a reduction in late-stage incidence and improved survival [5]. An explanation for the discrepancy is that achieving a mortality reduction requires a screening strategy capable of detecting OC even earlier and in a larger proportion of women than these trials were able to achieve [2]. While cancer mortality remains the key outcome, stage at diagnosis is considered a valuable surrogate endpoint, as it can be used to predict cancer death in various ways [38].

Few further study limitations should be noted. First, the reported accuracy of USS for OC detection, which relies heavily on sonographers’ expertise and experience, varies markedly between studies [16, 25, 41–43], with a lack of data from the primary care setting. For the base-case analysis, we adopted USS accuracy used in two UK OC clinical pathway evaluation studies [11, 16], where the accuracy value is about the average among previously reported data. We also explored varying USS accuracy in the sensitivity analysis. Second, the patients referred to secondary care who were ultimately not diagnosed with OC or any other cancers represent a heterogeneous group, and the treatment they received cannot be determined from the CPRD data. To address this, we made assumptions regarding the proportion undergoing surgery and the type of procedures, informed by consultations with our collaborating gynaecologists. Third, data on the long-term QoL impact of surgery for benign gynaecological disease is limited [22]. We made assumptions informed by the literature [22, 23] and conducted sensitivity analyses [44, 45]. Key study assumptions are listed in **Supplementary Table S10**.

This study highlights that using risk-based triage incorporating CA125 value and age to replace the current CA125 ≥ 35 U/mL threshold would be cost-effective among women aged over 50 years. Setting the moderate-risk threshold (for subsequent USS) at 1% and the high-risk threshold (for referral to secondary care) at 3% in Ovatools or using equivalent age-group specific CA125 thresholds, resulted in an ICER between £20,000 – £30,000. A major driver of the additional costs is the increased number of hospital referrals. Raising the threshold for subsequent USS significantly reduced the ICER. For example, setting the threshold for primary care USS at 1.2% or 1.5% Ovatools risk would achieve ICERs of approximately £20,000 and £10,000, respectively (**Supplementary Table S7 & S8**). As only age and the CA125 value are required to determine Ovatools risk, risk could readily be provided to clinicians with CA125 results in laboratory reports alongside clinical recommendations. Alternatively, simpler age-group specific CA125 thresholds, similar to those already use to interpret the prostate cancer test PSA in UK primary care [7], could be used in place of the single CA1245 cut-off, to guide clinical decision making.

Further research is needed to address the uncertainty in late-stage OC risk reduction due to earlier detection in primary care. Empirical data from trials or modelling studies using natural history models with estimations of dwell time in early stages would enhance understanding in this area. Additionally, studies that measure the long-term QoL impact of benign gynaecological surgery would contribute to better estimation of the benefits from early detection of gynaecological cancers, as these approaches are likely to increase hospital referrals for patients with only benign conditions. Further studies investigating the accuracy of USS for OC detection in primate care and, given accuracy is user dependent, the impact of changes in USS accuracy and costs on the cost-effectiveness would help ensure we are using optimal testing approaches.

Pathways with concurrent USS are not cost-effective due to high USS costs. However, increasing the use of USS in community diagnostic centres with sonographers with specific training who interpret scan in line with gold standard protocols [25] could enhance cost-effectiveness of these pathways. Finally, exploring the feasibility and acceptability of these new pathways among patients and clinicians could support effective implementation of a new pathway in primary care.

In conclusion, for women over 50 years of age, CA125-USS sequential pathways using Ovatools probabilities or equivalent age-adjusted CA125 thresholds and concurrent CA125-USS pathways improved OC detection with the sequential pathways potentially cost-effective. For women under 50 years, only pathways with concurrent CA125 and USS improved OC detection but were not cost-effective.

## Acknowledgements

We would like to thank Prof. Sudha Sundar, Dr. Samuel Perry, Dr. Mark Monahan and Mr. Donal Griffin at the University of Birmingham for their valuable data and support. We are also grateful to Prof. Bethany Shinkins at the University of Warwick for their insightful feedback and comments on this study.

This study is based in part on data from the Clinical Practice Research Datalink obtained under licence from the UK Medicines and Healthcare products Regulatory Agency. The data is provided by patients and collected by the NHS as part of their care and support. The interpretation and conclusions contained in this study are those of the author/s alone. The research has also used data from UK Biobank Resource under Application Number 56757 www.ukbiobank.ac.uk.

## Authors’ contributions

Conceptualisation: BM, FWM, GF & RW; Data curation: KDA; Analysis: RW, KDA & TH; Funding acquisition: RW, GF, BM & FWM; Investigation: RW, GF, BM, KDA, EJC & FWM; Methodology: RW & BM; Project management: RW & GF; Writing – original draft: RW; Writing – review and editing: RW, BM, GF, KDA, TH, FWM & EJC.

## Ethics

The study was approved by the Independent Scientific Advisory Committee (ISAC) for the Medicines and Healthcare Products Regulatory Agency (protocol number 21_001655). All data were provided to researchers in an anonymised form, and individual consent was not required.

## Data availability

The data used for this study were provided by CPRD and NCRAS and are subject to a licensing agreement that prohibits sharing outside the research team. Data can be requested through CPRD. Other data underlining this work may be obtained from third parties (UK Biobank: https://www.ukbiobank.ac.uk) and are not publicly available. Researchers can apply to use the UK Biobank resource. The R code of the health economic model is available at https://github.com/RunguoWu/Ovatools.

## Competing Interests

The authors declare no conflict of interest.

## Funding information

This study/project is funded by the National Institute for Health and Care Research (NIHR) School for Primary Care Research (project reference 629). EJC is supported by a National Institute for Health and Care Research (NIHR) Advanced Fellowship (NIHR300650) and the NIHR Manchester Biomedical Research Centre (NIHR203308). The views expressed are those of the author(s) and not necessarily those of the NIHR or the Department of Health and Social Care. This research arises from the CanTest Collaborative, funded by Cancer Research UK [C8640/A23385].

## Notes

### Competing Interest Statement

The authors have declared no competing interest.

### Author Declarations

The study used ONLY openly available human data that were originally located at CPRD, NCRAS and UK Biobank. Application is needed for the use of these data sources.

### Summary of Updates

I changed a co-author's first name from Boby to Borislava. Boby is her simplified informal name for daily use, but I used it here due to an oversight. Borislave is her name used in formal places.

