## Supplementary Materials for "Cost-effectiveness of CA125- and age-informed risk-based triage for ovarian cancer detection in primary care" (from another version of this preprint: version 1, 2025-04-01)

Cost-effectiveness analysis of implementing risk-based triage for ovarian cancer detection using Ovatools in UK primary care

**Supplementary materials**

### Supplementary Method 1: The pathway of CA125 with varying thresholds by age groups

In the CA125 *age-adjusted threshold sequential pathway*, age group-specific CA125 thresholds with accuracy equating to the accuracy of Ovatools 1% and 3% risk probabilities within age group were used in this study, with direct referral above the 3% threshold and pelvic USS for 1%-2.9%. The accuracy data was informed by CPRD Aurum validation data (**Table SM1**)^1^. In the analysis, we used the average accuracy data using this method across 18-49 and 50-89 to parametrize the decision tree, as we did with Ovatools accuracy data. See <https://qmro.qmul.ac.uk/xmlui/handle/123456789/105285> for further information.

**Table SM1: Accuracy of CA125 using age group-stratified levels equivalent to 1% and 3% Ovatools risk for detecting invasive ovarian cancer**

| Age group (invasive OC incidence) | % risk or CA125 equivalent threshold | Sensitivity, % (95% CI) | Specificity, % (95% CI) |
| --- | --- | --- | --- |
| All ages 18-89 years (0.62%) | ≥35 | 84.7 (83.1; 86.2) | 93.7 (93.6; 93.8) |
|  | applied by age category | 87.1 (85.5; 88.5) | 92.0 (91.9; 92.1) |
|  |  | 77.2 (75.4; 79.0) | 97.7 (97.6; 97.7) |
| All ages 18-49 years (0.21%) | ≥35 | 75.3 (70.0; 80.0) | 92.5 (92.3; 92.6) |
|  | Average accuracy, applied by age category | 63.5 (57.8; 69.0) | 96.9 (96.8; 96.9) |
|  |  | 43.5 (37.8; 49.3) | 99.3 (99.3; 99.4) |
| All ages 50-89 years (0.92%) | ≥35 | 86.2 (84.6; 87.8) | 94.6 (94.5; 94.7) |
|  | Average accuracy, applied by age category | 91.0 (89.6; 92.3) | 88.5 (88.3; 88.6) |
|  |  | 82.8 (81.0; 84.6) | 96.5 (96.4; 96.6) |
| 18-29 years (0.13%) | ≥35 | 56.5 (34.5; 76.8) | 94.8 (94.4; 95.1) |
|  | ≥34 (equating to 1%) | 56.5 (34.5; 76.8) | 94.4 (94.1; 94.8) |
|  | ≥91 (equating to 3%) | 30.4 (13.2; 52.9) | 98.8 (98.6; 98.9) |
| 30-39 years (0.14%) | ≥35 | 69.1 (55.2; 80.9) | 92.4 (92.1; 92.6) |
|  | ≥59 (equating to 1%) | 50.9 (37.1; 64.6) | 97.3 (97.1; 97.5) |
|  | ≥160 (equating to 3%) | 29.1 (17.6; 42.9) | 99.4 (99.3; 99.5) |
| 40-49 years (0.26%) | ≥35 | 78.7 (72.7; 83.9) | 92.0 (91.9; 92.2) |
|  | ≥58 (equating to 1%) | 67.4 (60.8; 73.6) | 97.2 (97.1; 97.3) |
|  | ≥157 (equating to 3%) | 48.4 (41.7; 55.2) | 99.4 (99.4; 99.5) |
| 50-59 years (0.52%) | ≥35 | 80.5 (76.3; 84.3) | 95.7 (76.3; 84.3) |
|  | ≥26 (equating to 1%) | 84.8 (80.9; 88.2) | 91.6 (91.4; 91.8) |
|  | ≥57 (equating to 3%) | 72.3 (67.7; 76.6) | 98.1 (98.0; 98.2) |
| 60-69 years (1.05%) | ≥35 | 86.9 (83.9; 89.5) | 95.9 (95.8; 96.1) |
|  | ≥22 (equating to 1%) | 92.4 (90.0; 94.4) | 89.3 (89.0; 89.5) |
|  | ≥37 (equating to 3%) | 86.6 (83.6; 89.2) | 96.2 (96.1; 96.4) |
| 70-79 years (1.32%) | ≥35 | 87.7 (84.6; 90.3) | 93.6 (93.4; 93.8) |
|  | ≥22 (equating to 1%) | 93.5 (91.0; 95.4) | 84.7 (84.4; 85.1) |
|  | ≥41 (equating to 3%) | 86.4 (83.2; 89.2) | 94.9 (94.6; 95.1) |
| 80-89 years (1.26%) | ≥35 | 90.6 (88.6; 93.9) | 88.6 (88.1; 89.0) |
|  | ≥26 (equating to 1%) | 92.2 (88.1; 95.1) | 81.8 (81.2; 82.3) |
|  | ≥58 (equating to 3%) | 83.1 (78.0; 87.5) | 94.0 (93.6; 94.3) |

### Supplementary Method 2: Estimation of cancer incidence and stage

Using the population from the Clinical Practice Research Datalink (CPRD), incident cancers within 1 year from the index date were identified, including ovarian cancer, lower gastrointestinal (GI) cancer, lung cancer, pancreatic cancer, uterine cancer and other cancers. Separate logistic regression models of incident cancer were fitted for different cancer types with 10-year age band, ethnicity and Townsend score quintiles as covariates. These models were then used to predict incidence of these cancers in these categories (**Table SM2 & SM3**).

Following missing data imputation, logistic regression models with the same specification were also estimated for cancer stage, i.e. early stage (stage I and II) or late stage (stage III and IV) and used to predict the proportion of late-stage cancer at diagnosis (**Table SM2 & SM3**).

In the base-case cost-effectiveness analysis, the pathway effects on non-ovarian cancers were not included, but their incidence was accounted for in modelling survival.

**Table SM2: The logistic regression models for cancer incidence and cancer stage, by cancer type, among women with CA125 records**

|  | Ovarian cancer | | Uterine cancer | |
| --- | --- | --- | --- | --- |
|  | Incidence | Late stage | Incidence | Late stage |
|  | Coefficient (Standard error) | | | |
| Intercept | -6.36 (0.13) | -0.61 (0.29) | -7.7 (0.16) | -1.13 (0.36) |
| Age 40-49 | 0.45 (0.14) | 0.55 (0.33) |  |  |
| Age 50-59 | 1.32 (0.13) | 0.98 (0.31) | 1.53 (0.16) | 0.07 (0.41) |
| Age 60-69 | 2.04 (0.13) | 1.48 (0.31) | 2.26 (0.15) | 0.22 (0.39) |
| Age 70-79 | 2.31 (0.13) | 1.63 (0.31) | 2.29 (0.15) | 0.56 (0.4) |
| Age >=80 | 2.18 (0.14) | 2.09 (0.34) | 2.24 (0.17) | 0.4 (0.43) |
| Asian | -0.2 (0.13) | 0.33 (0.3) | 0.17 (0.19) | -0.59 (0.55) |
| Black | -0.49 (0.2) | 0.68 (0.5) | 0.36 (0.22) | 1.45 (0.52) |
| Others | -0.67 (0.41) | -0.54 (1.02) | * | * |
| Townsend Q1 | -0.1 (0.07) | -0.09 (0.14) | 0.05 (0.12) | -0.18 (0.25) |
| Townsend Q2 | -0.07 (0.07) | -0.19 (0.13) | 0.24 (0.12) | -0.26 (0.22) |
| Townsend Q4 | 0 (0.07) | 0.13 (0.13) | 0.11 (0.13) | 0.04 (0.21) |
| Townsend Q5 | -0.26 (0.09) | -0.06 (0.11) | 0.03 (0.14) | 0.05 (0.19) |

**Table SM2: continued**

|  | Lung cancer | | Pancreatic cancer | |
| --- | --- | --- | --- | --- |
|  | Incidence | Late stage | Incidence | Late stage |
|  | Coefficient (Standard error) | | | |
| Intercept | -7.6 (0.14) | 1.27 (0.28) | -7.94 (0.16) | 1.68 (0.37) |
| Age 40-49 |  |  |  |  |
| Age 50-59 |  |  |  |  |
| Age 60-69 | 2.05 (0.14) | 0.11 (0.34) | 1.77 (0.16) | -0.28 (0.44) |
| Age 70-79 | 2.51 (0.13) | -0.43 (0.32) | 2.34 (0.15) | 0.14 (0.46) |
| Age >=80 | 2.86 (0.14) | 0.07 (0.34) | 2.89 (0.15) | 0.17 (0.46) |
| Asian | -1.71 (0.45) | -0.99 (1.01) | -1.19 (0.45) | 12.98 (627.24) |
| Black | -1.28 (0.45) | -1.49 (0.95) | -0.38 (0.39) | -0.7 (1.1) |
| Others | * | * | * | * |
| Townsend Q1 | -0.34 (0.13) | -0.42 (0.24) | 0.18 (0.14) | 0.18 (0.4) |
| Townsend Q2 | -0.22 (0.13) | 0.21 (0.25) | 0.02 (0.15) | 0.17 (0.37) |
| Townsend Q4 | 0.28 (0.13) | 0.11 (0.21) | 0.16 (0.16) | 0.15 (0.33) |
| Townsend Q5 | 0.32 (0.14) | 0.54 (0.23) | 0.17 (0.18) | 0.17 (0.31) |

**Table SM2: continued**

|  | Low GI cancer | | Other cancers | |
| --- | --- | --- | --- | --- |
|  | Incidence | Late stage | Incidence | Late stage |
|  | Coefficient (Standard error) | | | |
| Intercept | -6.92 (0.11) | 0.43 (0.22) | -5.75 (0.1) | -1.03 (0.23) |
| Age 40-49 |  |  | 0.62 (0.1) | 0.09 (0.27) |
| Age 50-59 | 1.23 (0.12) | 0.14 (0.26) | 1.06 (0.1) | 0.48 (0.26) |
| Age 60-69 | 1.9 (0.12) | 0.26 (0.25) | 1.65 (0.1) | 0.89 (0.24) |
| Age 70-79 | 2.5 (0.11) | 0.03 (0.24) | 2.02 (0.1) | 1.22 (0.24) |
| Age >=80 | 2.95 (0.11) | 0.17 (0.25) | 2.41 (0.1) | 1.43 (0.25) |
| Asian | -1.38 (0.28) | 0.1 (0.62) | -0.41 (0.11) | -0.44 (0.27) |
| Black | -0.78 (0.27) | -0.33 (0.59) | -0.43 (0.15) | 0.38 (0.35) |
| Others | * | * | -0.32 (0.27) | 0.43 (0.63) |
| Townsend Q1 | 0 (0.08) | -0.11 (0.15) | -0.08 (0.06) | 0.18 (0.11) |
| Townsend Q2 | -0.02 (0.08) | 0.11 (0.15) | -0.06 (0.06) | 0.02 (0.1) |
| Townsend Q4 | -0.02 (0.09) | -0.03 (0.14) | -0.02 (0.06) | 0.02 (0.1) |
| Townsend Q5 | 0.04 (0.1) | 0.13 (0.13) | -0.08 (0.07) | -0.1 (0.1) |

* Combined with White due to a limited number of observations for other ethnicity.

**Table SM3: The logistic regression models for cancer incidence and cancer stage, by cancer type, among women with CA125 or USS records**

|  | Ovarian cancer | | Uterine cancer | |
| --- | --- | --- | --- | --- |
|  | Incidence | Late stage | Incidence | Late stage |
|  | Coefficient (Standard error) | | | |
| Intercept | -6.96 (0.11) | -0.66 (0.25) | -7.84 (0.13) | -1.24 (0.3) |
| Age 40-49 | 0.88 (0.13) | 0.57 (0.29) |  |  |
| Age 50-59 | 1.8 (0.11) | 0.98 (0.28) | 1.81 (0.13) | 0.02 (0.35) |
| Age 60-69 | 2.55 (0.11) | 1.52 (0.27) | 2.6 (0.12) | 0.22 (0.32) |
| Age 70-79 | 2.77 (0.11) | 1.65 (0.28) | 2.76 (0.12) | 0.61 (0.32) |
| Age >=80 | 2.65 (0.12) | 1.96 (0.29) | 2.81 (0.13) | 0.73 (0.34) |
| Asian | -0.27 (0.11) | 0.3 (0.26) | 0.15 (0.14) | -0.04 (0.36) |
| Black | -0.65 (0.18) | 0.7 (0.47) | 0.21 (0.17) | 1.17 (0.39) |
| Others | -0.52 (0.34) | -0.06 (0.79) | * | * |
| Townsend Q1 | -0.06 (0.06) | -0.09 (0.14) | -0.01 (0.1) | -0.16 (0.19) |
| Townsend Q2 | -0.04 (0.06) | -0.21 (0.12) | 0.14 (0.09) | -0.19 (0.17) |
| Townsend Q4 | -0.02 (0.07) | 0.1 (0.13) | 0.05 (0.1) | 0.07 (0.17) |
| Townsend Q5 | -0.24 (0.08) | -0.1 (0.11) | 0.04 (0.11) | 0.04 (0.16) |

**Table SM3: continued**

|  | Lung cancer | | Pancreatic cancer | |
| --- | --- | --- | --- | --- |
|  | Incidence | Late stage | Incidence | Late stage |
|  | Coefficient (Standard error) | | | |
| Intercept | -7.88 (0.13) | 1.29 (0.26) | -8.14 (0.14) | 1.59 (0.31) |
| Age 40-49 |  |  |  |  |
| Age 50-59 |  |  |  |  |
| Age 60-69 | 2.36 (0.12) | 0.01 (0.31) | 2 (0.14) | -0.17 (0.39) |
| Age 70-79 | 2.75 (0.12) | -0.46 (0.29) | 2.56 (0.14) | 0.2 (0.4) |
| Age >=80 | 3.07 (0.13) | 0.03 (0.32) | 3.02 (0.14) | 0.32 (0.41) |
| Asian | -1.79 (0.38) | -0.36 (0.89) | -1.42 (0.41) | -0.09 (1.13) |
| Black | -1.53 (0.41) | -1.09 (0.85) | -0.45 (0.33) | -0.44 (1.05) |
| Others | * | * | * | * |
| Townsend Q1 | -0.34 (0.12) | -0.47 (0.22) | 0.14 (0.13) | 0 (0.34) |
| Townsend Q2 | -0.21 (0.12) | 0.32 (0.22) | -0.01 (0.14) | 0.21 (0.33) |
| Townsend Q4 | 0.23 (0.12) | 0.04 (0.2) | 0.11 (0.15) | 0.22 (0.3) |
| Townsend Q5 | 0.34 (0.12) | 0.48 (0.21) | 0.12 (0.16) | 0.21 (0.28) |

**Table SM3: continued**

|  | Low GI cancer | | Other cancers | |
| --- | --- | --- | --- | --- |
|  | Incidence | Late stage | Incidence | Late stage |
|  | Coefficient (Standard error) | | | |
| Intercept | -7.21 (0.1) | 0.37 (0.18) | -6.09 (0.07) | -0.96 (0.17) |
| Age 40-49 |  |  | 0.88 (0.08) | 0.01 (0.21) |
| Age 50-59 | 1.42 (0.1) | 0.19 (0.23) | 1.37 (0.08) | 0.39 (0.2) |
| Age 60-69 | 2.16 (0.1) | 0.4 (0.22) | 1.97 (0.08) | 0.78 (0.19) |
| Age 70-79 | 2.66 (0.1) | 0.07 (0.21) | 2.36 (0.08) | 1.11 (0.19) |
| Age >=80 | 3.11 (0.1) | 0.22 (0.21) | 2.67 (0.08) | 1.3 (0.19) |
| Asian | -1.05 (0.2) | 0.08 (0.43) | -0.48 (0.09) | -0.29 (0.22) |
| Black | -0.48 (0.19) | 0.16 (0.44) | -0.42 (0.11) | 0.23 (0.28) |
| Others | * | * | -0.41 (0.23) | 0.18 (0.54) |
| Townsend Q1 | 0.06 (0.08) | -0.13 (0.14) | -0.07 (0.05) | 0.1 (0.09) |
| Townsend Q2 | 0.05 (0.08) | 0.08 (0.13) | -0.07 (0.05) | 0 (0.09) |
| Townsend Q4 | 0.03 (0.09) | -0.03 (0.13) | 0.01 (0.06) | -0.04 (0.09) |
| Townsend Q5 | 0.07 (0.09) | 0.1 (0.12) | -0.05 (0.06) | -0.06 (0.09) |

* Combined with White due to a limited number of observations for other ethnicity.

### Supplementary Method 3: Estimation of stage shift parameters

We considered the potential shift in cancer stage at diagnosis for patients who were finally diagnosed at the late stage of cancer if they were in false negative in the original pathway but in true positive in the new pathway. A fraction of additionally detected ‘previously late stage’ cancers would ‘shift’ to an early stage at diagnosis in the new pathway. This fraction was calculated using data as **Table SM3** described, using the relative risk ratio of late-stage OC incidence, derived from data for screen detected cases and clinically detected cases among symptomatic women in UKCTOCS trial, where the screen detected cases were considered as earlier detection and the clinically detected cases were considered as usual care detection.^2^ We also considered the possibility of opposite direction of stage shift using the same method. We assumed cancer cases detected at an earlier stage to have their disease-specific life expectancy following that of an early-stage diagnosis and the survival origin for stage-shifted cases to be their original time of clinical diagnosis.^3^

**Table SM3: Relative risk ratio of diagnosis at the early/late stage between earlier-detected cases and general cases among symptomatic women**

|  | **Diagnosis at late stage** | **Data source** |
| --- | --- | --- |
| Ovarian cancer | 0.84 (0.74 – 0.95) | UKCTOCS^2^ |

The example of calculating relative risk ratio (RRR) of late-stage diagnosis between earlier detected ovarian cancers and other ovarian cancer cases for symptomatic women is presented as follows.

The UKCTOCS trial of ovarian cancer screening reported 159 early-stage cases and 602 late-stage cases in the non-screening-detected group, and 45 early-stage cases and 88 late-stage cases in the screening-detected group.

RRR = [88 / (88 + 45)] / [602 / (602 + 159)]

Var(ln (RRR)) = 1/88 – 1/(88 + 45) + 1/602 – 1/ (602 + 159)

### Supplementary Method 4: Addressing missing values for cancer stages

About 30% of cancer stage data from linked cancer registration data were missing. For estimating cancer stage distribution at diagnosis and fitting the survival models and costs models, we used multivariate imputation by chained equations (MICE) with a proportional odds logistic model in R to impute the missing values.^4^

In addition to all analytic covariates included in fitted models, follow-up time from diagnosis, ever died from cancer, ever died from other cause/s, ovarian-cancer-like symptoms and cancer behaviour (a variable of tumour nature, also from cancer registration data with missing values coded as a level, not imputed) were included as auxiliary variables in the multiple imputation. 30 imputed datasets were generated. Stages 1 to 4 were imputed separately and were then combined into early stage (stage I and II) and late stage (stage III and IV) for analysis. Results from the regression models fitted on each of the 30 datasets were combined using Rubin’s rule.^5^

### Supplementary Method 5: Cancer death risk estimation and model validation

Prediction models on death caused by cancer (death records with cancer as a cause in the CPRD linked death registration data) from the date of diagnosis were fitted based on patients with cancer diagnosed within 1 year following the index date using data for the CPRD patients with their linked Hospital Episode Statistics (HES) inpatient care and cancer registration data. The population include women with CA125 or USS records (with OC related symptoms for those who only have USS records). Separate models were fitted by different cancer types, including ovarian cancer, lung cancer, lower GI cancer, pancreatic cancer, uterus cancer, and other cancers combined as a whole. The date of diagnosis was defined as the earliest recorded date in hospital admissions and cancer registration with a diagnostic record of the corresponding cancer type.

The modelling method followed the cancer survival methodology used by the National Cancer Registration and Analysis Service (NCRAS).^6^ Flexible parametric models were fitted,^7^ with age at diagnosis, cancer stage of diagnosis (early/late), ethnicity (other ethnicity was combined with White as the category has too few observations for a few types of cancer) and socioeconomic deprivation level as the covariates, using the “rstpm2” package in R.^8^ Up to 5 degrees of freedom were allowed for both the baseline hazard function and time-dependent effects, with the best-fitted model selected by Akaike Information Criterion. The estimated models are presented in **Table SM5.1.**

These risk models were internally validated by plotting model predicted survival rates against observed survival trajectories up to 8 years of follow-up from diagnosis, for each modelled cancer type demonstrating good correspondence (**Figure SM5**). The model performance was also checked in women with CA125 records and women with only USS records, separately, and both demonstrated good fit. These risk models were integrated into the Markov model to predict cancer death.

Finally, the Markov model predictions for ovarian cancer, uterine cancer, lung cancer, lower GI cancer and pancreatic cancer mortality were externally validated against NHS England data using 5-year overall survival rates (**Table SM5.2**). The model predicted survival rates (weighted by stage distributions) were generally consistent, though slightly higher, with the NHS England data. A possible explanation is that the average age of the study population is 54.6 years, younger than the typical patient populations of cancers such as pancreatic cancer and lung cancer. For example, most pancreatic cancer happened among people aged over 75 years, who had a significantly lower survival rate than patients aged 50 with pancreatic cancer.

**Figure SM5: Plots of model fitted survival curves for death caused by cancer against observed data across cancer types in flexible parametric risk models**

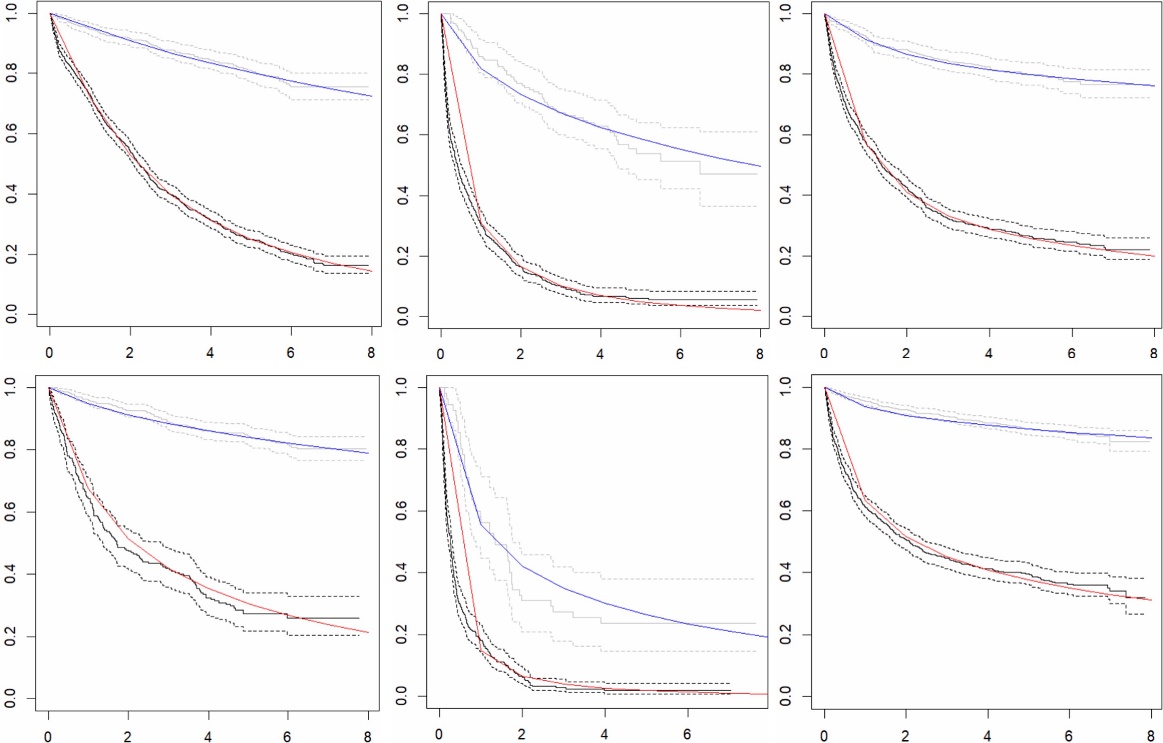

From left to right ovarian cancer, lung cancer, lower GI cancer in the first line, and uterine cancer, pancreatic cancer and other cancers in the second line. The blue lines indicate survivals with an early-stage diagnosis and the read lines indicate that with a late-stage diagnosis.

**Table SM5.1: Coefficients of flexible parametric models for cancer death by cancer types**

|  | **Ovarian** | **Lower GI** | **Lung** | **Pancreatic** | **Uterus** | **Others** |
| --- | --- | --- | --- | --- | --- | --- |
| Intercept | -7.64 (0.31) | -7.42 (0.34) | -7.46 (0.47) | -7.9 (0.44) | -8.71 (0.55) | -7.45 (0.24) |
| Age, centred at 60 divided by 10 | 0.41 (0.03) | 0.3 (0.03) | 0.21 (0.04) | 0.26 (0.04) | 0.43 (0.05) | 0.42 (0.02) |
| Asian ethnicity | -0.12 (0.18) | -0.66 (0.32) | -0.36 (0.47) | 0.01 (0.46) | -0.19 (0.32) | -0.07 (0.2) |
| Black ethnicity | 0.02 (0.27) | -0.21 (0.29) | -0.12 (0.51) | -0.44 (0.41) | 0.6 (0.24) | -0.24 (0.22) |
| Townsend score quintile Q1 (least deprived) | -0.05 (0.08) | -0.05 (0.1) | -0.18 (0.13) | -0.25 (0.14) | 0.31 (0.16) | -0.17 (0.09) |
| Townsend score quintile Q2 | 0 (0.09) | 0.01 (0.11) | -0.21 (0.13) | 0.01 (0.14) | -0.07 (0.17) | -0.02 (0.09) |
| Townsend score quintile Q4 | 0.06 (0.1) | 0.07 (0.12) | -0.31 (0.13) | 0.02 (0.16) | 0.06 (0.19) | -0.03 (0.1) |
| Townsend score quintile Q5 | 0.03 (0.11) | 0.16 (0.12) | -0.17 (0.14) | -0.02 (0.17) | 0.36 (0.2) | 0.05 (0.1) |
| Late-stage diagnosis | 1.73 (0.1) | 1.91 (0.11) | 1.78 (0.14) | 1.26 (0.17) | 1.83 (0.12) | 1.97 (0.08) |
| nsx(log(fu_diag2cens), df = n)1 | 4.35 (0.28) | 4.42 (0.3) | 5.23 (0.43) | 11.75 (0.72) | 9.95 (0.96) | 4.31 (0.22) |
| nsx(log(fu_diag2cens), df = n)2 | 5.12 (0.29) | 4.29 (0.21) | 4.81 (0.3) | 4.63 (0.18) | 4.02 (0.21) | 4.02 (0.16) |
| nsx(log(fu_diag2cens), df = n)3 | 4.6 (0.19) | 8.06 (0.6) | 10.05 (0.87) | NA | NA | 7.86 (0.44) |
| nsx(log(fu_diag2cens), df = n)4 | 8.92 (0.56) | 4.16 (0.15) | 4.76 (0.2) | NA | NA | 3.9 (0.11) |
| nsx(log(fu_diag2cens), df = n)5 | 4.46 (0.14) | NA | NA | NA | NA | NA |

**Table SM5.2: Comparison of model prediction with NHS England data for average 5-year overall survival rate**

|  | **Model predicted** | **NHS England data^9^ (non-standardised)** |
| --- | --- | --- |
| Ovarian | 58.8% | 56.2% |
| Uterine | 76.5% | 75.0% |
| Lung | 28.4% | 26.8% |
| Pancreatic | 16.0% | 10.0%^a^ |
| Lower GI | 58.8% | 55.0%^b^ |

a. The study population are significantly younger than typical population of pancreatic cancer. The 5-year overall survival rates for pancreatic cancer among 45-54, 55-64 and 65-74 are 24.9%, 17.2% and 10.7% respectively.
b. Weighted Combined bowel, colon and rectal cancers’ survival rates.

### Supplementary Method 6: Inpatient care cost models and assumptions

The CPRD linked HES inpatient data with Health Resource Group (HES HRG) codes were used to identify the hospital care costs. In the HES HRG data, core spell codes, or episode codes when core spell codes were missing for the whole spell, were used to calculate costs per admission (spell), by mapping them to NHS reference costs.^10^ The costing process started with mapping to NHS England reference costs 2021-22. Some HRG codes in the HES HRG data were only used in some particular years. If the HRG codes were not available in the 2021-22 reference costs, they were mapped to reference costs in previous years and so on until 2009-10. A few HRG codes not available in these reference costs but available in the NHS England national tariff ^11^ were mapped to the tariff prices using the same method above. Few hospital admissions with completely missing HRG codes were costed using the average cost estimated by the total cost divided by the total number of service activities in NHS England reference costs 2021-22. All costs were inflated to year 2022 using the NHS cost inflation index (for costs before 2016, a different inflation index was used as NHS changed the rule of calculation in 2016).^12 13^

The statistical analysis followed previously published methods of estimating prediction models for health care costs.^14 15^ Separate models were fitted by cancer type, including ovarian cancer, lung cancer, lower GI cancer, pancreatic cancer, uterine cancer, and other cancers combined as a whole, and those without cancer diagnosis. Overlaps between cancer types were rare so their effects were not considered. Annual hospital care costs were calculated over annual periods from the date of diagnosis by summing up the costs incurred by each patient during each annual period of follow-up. For the hospital admissions that spanned more than one year of follow-up, a share of the total cost was allocated into each respective annual period based on the duration of the admission in the annual period. The cost model included annually updated patient age and durations from first diagnoses of the modelled cancer type (from the index date in the non-cancer model). For each disease, duration since the first diagnosis was defined as a categorical variable with the following levels: years before diagnosis, year of diagnosis, year post diagnosis, two years post diagnosis, three years post diagnosis and four or more years post diagnosis. Patients with cancer history before the index date were not considered to have incident cancer of the corresponding type. Cancer stage at diagnosis (early/late) was included in the cost model as a covariate. Patients’ ethnicity and Townsend deprivation index of their residential Lower Layer Super Output Area (LSOA) were also considered in the cost model. Interaction between cancer stage at diagnosis and duration of cancer were overall significant in at least one part of all the two-part models and thus were included. Missing cancer stage data were multiply imputed using the same method as described in **Supplementary Method 4**.

Annual hospital care costs were modelled using a two-part model, due to a large number of zero costs, with the first part logistic regression modelling of probability of having any cost and the second part generalised linear regression model (GLM) on costs conditional on having incurred costs. Six different GLMs of three distributions (Gaussian, Poisson and Gamma) and two links (identity and natural log) were tested using modified Park’s test and the Hosmer-Lemeshow test for the second part of the two-part model. Cluster robust standard errors were used due to the lack of independence between annual periods for the same patient. **Table SM6** presents results of the cost models based on multiple imputation data as described previously.

Additionally, in primary care, we assumed that the CA125 test required 10 minutes of nurse time and that a further GP consultation occurred along with referral to secondary care. After referrals to hospital, we assumed that the surgery rate applied only to those with a positive USS result in the hospital USS check, with the probability estimated based on the same USS accuracy data.

**Table SM6: Coefficients of Two-Part models for hospital costs by cancer types and non-cancer**

|  | **Ovarian** | | **Lower GI** | | **Lung** | | **Pancreatic** | |
| --- | --- | --- | --- | --- | --- | --- | --- | --- |
| Two parts | P1 | P2 | P1 | P2 | P1 | P2 | P1 | P2 |
| Model types | Logistic | Poison-Log | Logistic | Poison-Log | Logistic | Poison-Log | Logistic | Poison-Log |
| Intercept | -0.23 (0.09) | 8.09 (0.05) | 0.11 (0.08) | 7.94 (0.03) | 0.54 (0.11) | 8.19 (0.04) | 0.77 (0.22) | 8.26 (0.06) |
| Age, centred at 60 divided by 10 | 0.11 (0.02) | -0.08 (0) | 0 (0.02) | -0.06 (0) | -0.05 (0.03) | -0.07 (0) | -0.09 (0.04) | -0.09 (0) |
| Asian ethnicity | -0.18 (0.11) | 0.05 (0.01) | 0.24 (0.13) | 0.08 (0.01) | 0.09 (0.24) | -0.1 (0) | 1.53 (0.49) | 0.13 (0.02) |
| Black ethnicity | 0.06 (0.15) | 0.12 (0.01) | 0.32 (0.16) | -0.12 (0.01) | -0.12 (0.19) | 0.06 (0) | -0.07 (0.28) | 0.01 (0.01) |
| Townsend score quintile Q1 (least deprived) | -0.08 (0.06) | 0.06 (0.01) | -0.09 (0.06) | -0.03 (0) | 0 (0.1) | 0.03 (0) | -0.21 (0.16) | -0.09 (0) |
| Townsend score quintile Q2 | -0.09 (0.07) | 0.04 (0) | -0.03 (0.07) | -0.01 (0) | -0.04 (0.09) | 0.04 (0) | -0.08 (0.17) | -0.16 (0.01) |
| Townsend score quintile Q4 | -0.04 (0.07) | 0.05 (0) | 0.17 (0.07) | 0.07 (0) | 0.03 (0.09) | 0.05 (0) | 0.04 (0.18) | -0.16 (0) |
| Townsend score quintile Q5 | 0.14 (0.08) | 0.24 (0) | 0.15 (0.08) | 0.09 (0) | 0.17 (0.09) | 0.21 (0) | -0.19 (0.19) | 0 (0) |
| Late-stage diagnosis | 0.06 (0.1) | 0.04 (0.08) | -0.06 (0.09) | 0.22 (0.05) | -0.28 (0.11) | 0.07 (0.06) | -0.16 (0.21) | 0.1 (0.08) |
| Diagnosis within 1 year | 4.11 (0.26) | 1.07 (0.05) | 4.85 (0.39) | 1.33 (0.03) | 2.61 (0.22) | 1.1 (0.04) | 3.06 (0.62) | 1.4 (0.07) |
| Diagnosis 1 year ago | -0.3 (0.11) | 0.39 (0.06) | 0.46 (0.09) | 0.32 (0.03) | -0.35 (0.13) | 0.29 (0.05) | -0.6 (0.32) | 0.43 (0.11) |
| Diagnosis 2 years ago | -0.44 (0.11) | 0.42 (0.06) | -0.29 (0.1) | 0.36 (0.03) | -0.48 (0.14) | 0.32 (0.04) | -1.04 (0.34) | 0.57 (0.08) |
| Diagnosis 3 years ago | -0.6 (0.12) | 0.6 (0.09) | -0.36 (0.1) | 0.33 (0.04) | -0.62 (0.16) | 0.34 (0.04) | -1.41 (0.4) | 0.85 (0.07) |
| Diagnosis over 4 years ago | -0.62 (0.11) | 0.57 (0.11) | -0.32 (0.1) | 0.5 (0.04) | -0.91 (0.18) | 0.27 (0.04) | -0.81 (0.36) | 0.76 (0.07) |
| Cancer death | 2.39 (0.12) | 0.7 (0.01) | 1.62 (0.12) | 0.47 (0) | 1.89 (0.14) | 0.34 (0) | 1.81 (0.29) | 0.45 (0.01) |
| Non-cancer death | 2.54 (0.41) | 0.46 (0.03) | 2.06 (0.32) | 0.75 (0.01) | 2.16 (0.36) | 0.51 (0.01) | 16.63 (367.68) | 0.43 (0.03) |
| Proportion of lost follow-up in this year | -2.09 (0.14) | -1 (0) | -1.84 (0.12) | -0.97 (0) | -1.76 (0.16) | -0.88 (0.01) | -2.38 (0.39) | -1.2 (0.02) |
| Late stge* Diagnosis within 1 year | -0.73 (0.31) | 0.21 (0.07) | -1.81 (0.43) | -0.05 (0.05) | 0.21 (0.27) | -0.17 (0.05) | -0.42 (0.68) | -0.21 (0.09) |
| Late stge* Diagnosis 1 year ago | 0.96 (0.15) | 0.18 (0.08) | 0.34 (0.14) | 0.29 (0.05) | 0.51 (0.19) | 0.03 (0.07) | 0.93 (0.44) | 0.65 (0.14) |
| Late stge* Diagnosis 2 years ago | 0.93 (0.15) | 0.08 (0.09) | 0.26 (0.14) | 0.23 (0.05) | 0.45 (0.22) | 0.09 (0.07) | 0.76 (0.53) | -0.03 (0.15) |
| Late stge* Diagnosis 3 years ago | 0.94 (0.16) | -0.02 (0.14) | 0.07 (0.16) | 0.18 (0.06) | 0.26 (0.28) | 0.1 (0.07) | 0.77 (0.69) | -0.61 (0.16) |
| Late stge* Diagnosis over 4 years ago | 0.76 (0.15) | 0.13 (0.18) | -0.15 (0.15) | -0.12 (0.07) | 0.13 (0.32) | 0.2 (0.07) | 0.12 (0.74) | -0.52 (0.22) |

**Table SM6: continued**

|  | **Uterus** | | **Other cancers** | | **Non-cancer** | |
| --- | --- | --- | --- | --- | --- | --- |
| Two parts | P1 | P2 | P1 | P2 | P1 | P2 |
| Model types | Logistic | Poison-Log | Logistic | Poison-Log | Logistic | Gamma-Log |
| Intercept | -0.44 (0.08) | 8.09 (0.02) | -0.42 (0.03) | 8.08 (0.01) | -0.97 (0) | 7.91 (0.01) |
| Age, centred at 60 divided by 10 | 0.12 (0.02) | 0 (0) | 0.09 (0.01) | -0.03 (0) | 0.11 (0) | 0.04 (0) |
| Asian ethnicity | -0.12 (0.12) | -0.02 (0.01) | -0.06 (0.05) | 0.09 (0) | -0.08 (0.01) | -0.06 (0.01) |
| Black ethnicity | 0.1 (0.16) | 0.62 (0.01) | 0.09 (0.07) | -0.02 (0.01) | -0.16 (0.01) | -0.04 (0.01) |
| Townsend score quintile Q1 (least deprived) | -0.08 (0.08) | 0.01 (0) | -0.19 (0.03) | -0.11 (0) | -0.2 (0) | -0.07 (0.01) |
| Townsend score quintile Q2 | 0.12 (0.08) | 0.03 (0) | -0.1 (0.03) | -0.08 (0) | -0.1 (0) | -0.03 (0.01) |
| Townsend score quintile Q4 | 0.07 (0.09) | 0.13 (0) | 0.05 (0.04) | -0.07 (0) | 0.09 (0) | 0.03 (0.01) |
| Townsend score quintile Q5 | 0.12 (0.09) | -0.02 (0) | 0.12 (0.04) | 0.03 (0) | 0.13 (0) | 0.05 (0.01) |
| Late-stage diagnosis | 0.21 (0.12) | 0.16 (0.07) | 0.24 (0.05) | 0.04 (0.02) |  |  |
| Diagnosis within 1 year | 4.11 (0.2) | 0.85 (0.03) | 3.72 (0.07) | 0.9 (0.01) | 0.54 (0) | 0.05 (0.01) |
| Diagnosis 1 year ago | -0.31 (0.09) | 0.2 (0.02) | 0.27 (0.04) | 0.31 (0.03) | 0.14 (0.01) | 0.14 (0.01) |
| Diagnosis 2 years ago | -0.49 (0.09) | 0.33 (0.03) | 0.02 (0.04) | 0.34 (0.03) | 0.05 (0.01) | 0.18 (0.01) |
| Diagnosis 3 years ago | -0.53 (0.1) | 0.19 (0.02) | -0.17 (0.05) | 0.34 (0.02) | 0 (0.01) | 0.2 (0.01) |
| Diagnosis over 4 years ago | -0.54 (0.09) | 0.14 (0.03) | -0.34 (0.05) | 0.34 (0.02) | -0.07 (0) | 0.22 (0.01) |
| Cancer death | 2.55 (0.18) | 0.57 (0.01) | 2 (0.08) | 0.69 (0.01) | 3.62 (0.04) | 1.07 (0.02) |
| Non-cancer death | 2.16 (0.35) | 0.6 (0.01) | 2.12 (0.15) | 0.82 (0.01) | 2.84 (0.02) | 0.84 (0.02) |
| Proportion of lost follow-up in this year | -1.73 (0.16) | -0.62 (0.01) | -1.59 (0.06) | -0.85 (0) | -1.99 (0.01) | -0.35 (0.02) |
| Late stge* Diagnosis within 1 year | -1 (0.37) | -0.02 (0.08) | -0.72 (0.13) | 0.13 (0.02) |  |  |
| Late stge* Diagnosis 1 year ago | 0.26 (0.19) | 0.09 (0.08) | 0.13 (0.08) | 0.41 (0.09) |  |  |
| Late stge* Diagnosis 2 years ago | 0.46 (0.23) | 0.16 (0.09) | 0.13 (0.09) | 0.29 (0.08) |  |  |
| Late stge* Diagnosis 3 years ago | 0.18 (0.25) | 0.09 (0.07) | 0.06 (0.1) | 0.29 (0.05) |  |  |
| Late stge* Diagnosis over 4 years ago | 0.16 (0.23) | 0.02 (0.1) | 0.03 (0.1) | 0.17 (0.06) |  |  |

### Supplementary Method 7: UK Biobank data for QoL modelling

The quality of life (QoL) model used data from 167,191 UK Biobank participants who completed the EuroQoL-5 Dimension (EQ-5D-5L) questionnaire in the 2019 survey. The UK Biobank is a large population cohort of 500,000 participants recruited between 2006 and 2010 across the UK and under continuous follow-up and re-surveys.^16^ HES and cancer registry records linked with the UK Biobank were employed to identify cancer diagnoses and dates. Additionally, HES data, UK Biobank baseline survey responses, and general practice Read codes were used to identify other disease histories.

The dependent variable, the QoL utility index, was derived from the five responses on the EQ-5D-5L questionnaire, assessing mobility, self-care, usual activities, pain and anxiety, using the NICE DSU value set.^17^ The QoL utility values ranged from −0.532 for the worst health state to 1 for full health, where 0 represents a health state equivalent to death and higher values indicate better QoL. Key independent variables included histories of ovarian cancer, lower GI cancer, upper GI cancer (including pancreatic, liver and biliary cancers), lung cancer, and uterine cancer, with the time intervals between diagnosis and the survey. The variable of upper GI was used to replace pancreatic cancer due to the limited number of identified pancreatic cancer diagnoses, and evidence suggesting similar QoL scores among pancreatic cancer patients and other upper GI cancer patients.^18^ Other independent variables included history of other cancers, age at survey, sex, ethnicity, Townsend scores quintiles. A small number of values for ethnicity and Townsend scores were missing. Missing ethnicities were imputed as white, the majority ethnicity. A small number of missing Townsend scores were imputed by a linear regression model on IMD scores (index of multiple deprivation), years and sources (England, Wales or Scotland) if IMD scores were available, and imputed by looking up an average Townsend score according to the rounded ordnance survey coordinates if IMD scores are missing, or imputed by looking up an average Townsend score for the areas of the assessment centres if IMD scores and ordnance survey coordinates were both missing.

A linear regression model was employed to estimate the QoL utility based on both women and men, in order to increase the number of cancer cases. Ordinary linear regression remains a popular choice for modelling EQ-5D utility, allowing for direct estimation of QoL changes associated with individual characteristics and disease events. Given the non-linear effect of age on EQ-5D utility, age splines with a knot at 70 years were utilized. Variables of ovarian cancer, upper GI cancer, lower GI cancer, uterus cancer and lung cancer were categorised into four levels initially: absence of such cancer, cancer diagnosed within the past year, cancer diagnosed 2-5 years ago, and cancer diagnosed over 5 years ago (**Table SM7.1**). In the modelling, the cancer variables were consolidated into three levels: absence of such cancer, cancer diagnosed within the past year and cancer diagnosed over 1 years ago, because the initial examination suggested that the levels of cancer diagnosed 2-5 years ago and cancer diagnosed over 5 years ago could be combined into one level, as there was no statistically significant difference between them in the QoL model. Uterine cancer was combined with ovarian cancer, due to the limited number of observations. The binary variables of other cancers and benign gynae diseases were incorporated. The model is presented in **Table SM7.2**.

**Table SM7.1: Characteristics of UKB participant contributing to QoL model**

|  | Overall |
| --- | --- |
| n | 167191 |
| QoL utility(mean (SD)) | 0.83 (0.17) |
| Age at survey (mean (SD)) | 66.60 (7.68) |
| Ethnicity (%) | |
| White | 162322 (97.1) |
| Asian | 1862 (1.1) |
| Black | 1226 (0.7) |
| Others | 1781 (1.1) |
| Townsend Score (%) | |
| Q1 | 68169 (40.8) |
| Q2 | 34585 (20.7) |
| Q3 | 27390 (16.4) |
| Q4 | 22452 (13.4) |
| Q5 | 14595 (8.7) |
| Ovarian cancer (%) | |
| No such cancer | 166796 (99.8) |
| <=1 yr | 39 (0.0) |
| 2-5 yrs | 96 (0.1) |
| >5 yrs | 260 (0.2) |
| Upper GI cancer (%) | |
| No such cancer | 166924 (99.8) |
| <=1 yr | 110 (0.1) |
| 2-5 yrs | 93 (0.1) |
| >5 yrs | 64 (0.0) |
| Lung cancer (%) | |
| No such cancer | 166945 (99.9) |
| <=1 yr | 92 (0.1) |
| 2-5 yrs | 116 (0.1) |
| >5 yrs | 38 (0.0) |
| Uterine cancer (%) |  |
| No such cancer | 166834 (99.8) |
| <=1 yr | 70 (0.0) |
| 2-5 yrs | 144 (0.1) |
| >5 yrs | 143 (0.1) |
| Lower GI cancer (%) |  |
| No such cancer | 166018 (99.3) |
| <=1 yr | 219 (0.1) |
| 2-5 yrs | 516 (0.3) |
| >5 yrs | 438 (0.3) |
| With other cancer | 9252 (5.5) |
| With benign gynae disease | 2884 (1.7) |

**Table SM7.2: QoL model: linear regression model**

| **Variable** | **Coefficient (95% CI)** |
| --- | --- |
| Intercept | 0.839 (0.838 , 0.841) |
| lspline(age_cent60, 1, marginal = F)1 | 0 (-0.001 , 0.002) |
| lspline(age_cent60, 1, marginal = F)2 | -0.035 (-0.039 , -0.032) |
| Townsend score (Reference = Q1) |  |
| Q2 | -0.008 (-0.01 , -0.006) |
| Q3 | -0.015 (-0.017 , -0.013) |
| Q4 | -0.027 (-0.029 , -0.024) |
| Q5 | -0.053 (-0.056 , -0.05) |
| Ethnicity (Reference = White) |  |
| Asian | 0.005 (-0.003 , 0.013) |
| Black | -0.013 (-0.023 , -0.004) |
| Others | -0.011 (-0.019 , -0.003) |
| Gynae cancer diagnosed within 1 year prior to EQ-5D measurement | -0.054 (-0.086 , -0.022) |
| Gynae cancer diagnosed at least 1 year prior to EQ-5D measurement | -0.013 (-0.026 , 0.001) |
| Upper GI cancer diagnosed within 1 year prior to EQ-5D measurement | -0.079 (-0.111 , -0.048) |
| Upper GI cancer diagnosed at least 1 year prior to EQ-5D measurement | -0.036 (-0.063 , -0.01) |
| Lung cancer diagnosed within 1 year prior to EQ-5D measurement | -0.11 (-0.144 , -0.076) |
| Lung cancer diagnosed at least 1 year prior to EQ-5D measurement | -0.065 (-0.092 , -0.039) |
| Lower GI cancer diagnosed within 1 year prior to EQ-5D measurement | -0.016 (-0.038 , 0.006) |
| Lower GI cancer diagnosed at least 1 year prior to EQ-5D measurement | -0.015 (-0.026 , -0.004) |
| Other cancers, all years | -0.022 (-0.026 , -0.018) |
| Benign gynae disease, all years | -0.034 (-0.04 , -0.027) |

Impact of age on QoL is predicted as follows: when age < 70, QoL=Age spline 1 * (age - 60)/10; when age >= 70, QoL=Age spline 1 + Age spline 2 * [(age - 60)/10 – 1].

The comparator is White woman, aged 60 years, at a mid-level of socioeconomic status without any cancer and gynae disease.

### Supplementary Method 8: Parameter uncertainty

In the probabilistic sensitivity analysis, we created 1000 sets of model parameters and used them for execute simulations to assess joint parameter uncertainty. These parameter sets were generated by the bootstrap method or Monte Carlos simulations. The following table summarises the methods used to generate the parameter uncertainties for probabilistic sensitivity analysis.

**Table SM8: Methods used to generate parameter uncertainties**

| **Parameters** | **Sources** | **methods** |
| --- | --- | --- |
| Test/scan accuracy data and benign disease surgery rate | External sources | Random sample from beta distributions |
| Diagnostic costs and costs related to false positive | NHS cost references | No uncertainty |
| Cancer treatment and long-term costs | Model estimated using CPRD and linkage data | Bootstrap sampling |
| Long-term QoL utility | Model estimated using UK Biobank | Bootstrap sampling |
| Model estimated cancer death risk | Model estimated using CPRD and linkage data | Bootstrap sampling |
| Utility/disutility due to benign gynaecological surgery and additional QoL decrement due to late-stage diagnosis | External sources | Random sampling from normal distributions |
| Relative risk ratio of diagnosis at late/early stage | External sources | Random sampling from Log-normal distribution |
| Cancer incidence and stage distribution | Model estimated using CPRD and linkage data | Bootstrap sampling |

### Supplementary Method 9: The scenario considering the effect of diagnostic pathway on other cancers

In the decision tree, while ovarian cancer enters either true positive (TP) or false negative (FN). Benign gynaecological disease and other cancers enter either false positive (FP) or true negative (TN). In a scenario analysis, we further consider the effect of pathway on lower GI cancer, lung cancer, pancreatic cancer and uterine cancer, because they are likely to be picked up by CA125 and a few they by USS among patients with ovarian cancer related symptoms.^19^ Distributions of these in FP and TN were informed by previous studies examining other cancers in the FP group of ovarian cancer detection (**Table SM9.1**).^19 20^

**Table SM9.1: Decision tree parameters for uterine, lower GI, lung and pancreatic cancers**

| **Parameters** | **Lower GI** | **Uterine** | **Lung** | **Pancreatic** |
| --- | --- | --- | --- | --- |
| **Uterine cancer** |  |  |  |  |
| Percentage of being in false positive of OC detection (with USS) ^20^ | 0.002 | 0.012 | 0 | 0 |
| Percentage of being in false positive of OC detection (with Ovatools 3%) ^19^ | 0.019 | 0.015 | 0.016 | 0.015 |

We considered the potential shift in cancer stage of diagnosis for patients who were finally diagnosed at the late stage of these cancer if they were in TN in the original pathway but in FP in the new pathway with the Ovatools involved. A fraction of additionally detected ‘previously late stage’ cancers would ‘shift’ to an early stage at diagnosis in the new pathway. This fraction was calculated using data as **Table SM9.2** described, by deriving the relative risk ratio of early/late-stage diagnosis between early detected cases and other cases, using the method as **Supplementary Method 3** has described.

We assumed that these non-ovarian cancers referred to secondary care by the pathways incurred an additional cost of £117 for the CT scan before diagnosis and treatment.^21^

**Table SM9.2: relative risk ratio of diagnosis at the early/late stage between earlier-detected cases and general cases**

|  | **Diagnosis at late stage** | **Data source** |
| --- | --- | --- |
| Lung cancer | 0.63 (0.55 – 0.72) | PLCO^22^ |
| Colorectal cancer | 0.85 (0.74 – 0.98) | A cohort study^23^ |
| Uterine cancer | 0.88 (0.78 – 1.00) | Based on ovarian cancer data above, adjusted by the ratio of dwelling time at the early stage of ovarian cancer and uterus cancer^24^ |
| Pancreatic cancer | 0.48 (0.27 – 0.85) | A cohort study^25^ |

### Supplementary Method 10: CA125 and Ovatools accuracy parameters

Accuracy of CA125 and Ovatools varies not only by age groups but also by cancer stage at diagnosis. **Table SM10.1** presents the by-age-and-stage accuracy data for CA125 and Ovatools, informed by our parallel study analysing CPRD Aurum data. In the base-case analysis, we used the by-age accuracy data, considering the by-stage data may not be accurate with the assumption of stage shift. However, it is still worthwhile to use the by-age-and-stage accuracy data for a sensitivity analysis.

**Table SM10.1: Accuracy of CA125 >35U/ml and Ovatools at >1% and >3% to detect invasive ovarian cancer, by age and stage**

| Sub-group | Outcome (incidence) | CA125 or Ovatools threshold | Sensitivity, % (95% CI) | Specificity, % (95% CI) |
| --- | --- | --- | --- | --- |
| 18-49 years | Early-stage invasive ovarian cancer (n=139, 0.10%) | CA125 ≥35U/mL | 63.3 (54.7; 71.3) | 92.5 (92.4; 92.7) |
|  |  | ≥1% | 43.9 (35.5; 52.5) | 97.0 (96.9; 97.1) |
|  |  | ≥3% | 24.5 (17.6; 32.5) | 99.4 (99.3; 99.4) |
|  | Late-stage invasive ovarian cancer (n=124, 0.09%) | CA125 ≥35U/mL | 92.7 (86.7; 96.6) | 92.5 (92.4; 92.7) |
|  |  | ≥1% | 84.7 (77.1; 90.5) | 97.0 (96.9; 97.1) |
|  |  | ≥3% | 69.4 (60.4; 77.3) | 99.4 (99.3; 99.4) |
| ≥50 years | Early-stage invasive ovarian cancer (n=441, 0.22%) | CA125 ≥35U/mL | 67.8 (63.2; 72.1) | 94.4 (94.3; 94.5) |
|  |  | ≥1% | 79.1 (75.0; 82.8) | 89.2 (89.0; 89.3) |
|  |  | ≥3% | 60.3 (55.6; 64.9) | 96.6 (96.5; 96.7) |
|  | Late-stage invasive ovarian cancer (n=1,123, 0.57%) | CA125 ≥35U/mL | 93.4 (91.8; 94.8) | 94.4 (94.3; 94.5) |
|  |  | ≥1% | 95.7 (94.4; 96.8) | 89.2 (89.0; 89.3) |
|  |  | ≥3% | 91.8 (90.0; 93.3) | 96.6 (96.5; 96.7) |

### Supplementary Method 11: Estimating ICERs with Ovatools accuracy at different thresholds

We applied different thresholds for the moderate risk and the high risk in the Ovatools-USS sequential pathway to replace the original 1% and 3% in the base case analysis and estimate the incremental cost-effectiveness ratios (ICERs). In this analysis, the moderate risk threshold ranges from 0.6% to 2% and the high risk threshold ranges from 2% to 4%. The Ovatools accuracy at these thresholds are presented in the **Table SM11.1**, informed by our parallel study analysing CPRD Aurum data. The analysis focused on women aged over 50 years, because among women aged under 50 years, the OC prevalence is so low that the Ovatools risk threshold has to reduce to a very low level to match the CA125 ≥ 35 U/mL threshold and the Ovatools do not show significant advantage over the CA125 ≥ 35 U/mL threshold (**Table SM11.2**).

We could also apply different CA125 thresholds for the moderate risk and the high risk in the age-adjusted sequential pathway, matching the Ovatools risk thresholds on accuracy. The result would be similar to the Ovatools result.

**Table SM11.1: Ovatools by-age accuracy at different risk thresholds**

|  | Under 50 years | | Over 50 years | |
| --- | --- | --- | --- | --- |
| Risk threshold | Sensitivity | Specificity | Sensitivity | Specificity |
| Ovatools ≥0.6% | 71.9 (66.4; 76.9) | 93.1 (93.0; 93.2) | 92.8 (91.6; 94.0) | 84.6 (94.4; 94.7) |
| Ovatools ≥0.8% | 66.9 (61.2; 72.2) | 95.6 (95.5; 95.7) | 91.9 (90.5; 93.1) | 87.2 (87.1; 87.4) |
| Ovatools ≥1% | 61.9 (56.1; 67.4) | 96.9 (96.8; 97.0) | 91.1 (89.7; 92.4) | 89.1 (88.9; 89.2) |
| Ovatools ≥1.2% | 60.2 (54.4; 65.8) | 97.7 (97.6; 97.8) | 90.6 (89.1; 91.9) | 90.6 (90.5; 90.7) |
| Ovatools ≥1.4% | 57.9 (52.0; 63.5) | 98.2 (98.1; 98.2) | 89.7 (88.2; 91.0) | 91.9 (91.7; 92.0) |
| Ovatools ≥1.6% | 57.2 (51.4; 62.9) | 98.5 (98.4; 98.6) | 89.1 (87.6; 90.5) | 92.9 (92.8; 93.0) |
| Ovatools ≥1.8% | 54.8 (49.0; 60.6) | 98.7 (98.7; 98.8) | 88.2 (86.7; 89.7) | 93.7 (93.6; 93.8) |
| Ovatools ≥2% | 52.8 (47.0; 58.6) | 98.9 (98.8; 98.9) | 87.4 (85.8; 88.8) | 94.4 (94.3; 94.5) |
| Ovatools ≥2.2% | 51.2 (45.4; 57.0) | 99.0 (98.9; 99.0) | 86.0 (84.3; 87.6) | 95.0 (94.9; 95.1) |
| Ovatools ≥2.4% | 49.2 (43.4; 55.0) | 99.1 (99.0; 99.1) | 85.1 (83.4; 86.7) | 95.5 (95.4; 95.6) |
| Ovatools ≥2.6% | 48.5 (42.7; 54.3) | 99.2 (99.2; 99.2) | 84.3 (82.5; 85.9) | 95.9 (95.8; 96.0) |
| Ovatools ≥2.8% | 46.8 (41.1; 52.7) | 99.3 (99.2; 99.3) | 83.6 (81.8; 85.2) | 96.2 (96.1; 96.3) |
| Ovatools ≥3% | 45.2 (39.4; 51.0) | 99.4 (99.3; 99.4) | 83.1 (81.3; 84.8) | 96.5 (96.4; 96.6) |
| Ovatools ≥3.2% | 43.8 (38.1; 49.6) | 99.4 (99.4; 99.4) | 81.9 (80.1; 83.7) | 96.8 (96.7; 96.9) |
| Ovatools ≥3.4% | 42.1 (36.5; 48.0) | 99.4 (99.4; 99.5) | 81.0 (79.1; 82.8) | 97.0 (96.9; 97.1) |
| Ovatools ≥3.6% | 41.5 (35.8; 47.3) | 99.5 (99.4; 99.5) | 80.2 (78.4; 82.0) | 97.2 (97.1; 97.2) |
| Ovatools ≥3.8% | 39.1 (33.6; 44.9) | 99.5 (99.5; 99.6) | 79.7 (77.8; 81.5) | 97.3 (97.3; 97.4) |
| Ovatools ≥4% | 38.5 (32.9; 44.2) | 99.6 (99.5; 99.6) | 78.9 (77.0; 80.8) | 97.5 (97.4; 97.5) |

**Table SM11.1: The Ovatools risk thresholds matched on CA125 ≥ 35 U/mL by age groups**

| Age group | Matched on | Threshold | Sensitivity | Specificity |
| --- | --- | --- | --- | --- |
| 18-29 years |  | CA125 >35 | 56.5 (34.5; 76.8) | 94.8 (94.4; 95.1) |
|  | sensitivity | Ovatools ≥0.8% | 56.5 (34.5; 76.8) | 93.6 (93.3; 94.0) |
|  | specificity | Ovatools ≥0.92% | 47.8 (26.8; 69.4) | 94.8 (94.5; 95.1) |
| 30-39 years |  | CA125 >35 | 69.1 (55.2; 80.9) | 92.4 (92.1; 92.6) |
|  | sensitivity | Ovatools ≥0.514% | 69.1 (55.2; 80.9) | 92.4 (92.2; 92.7) |
|  | specificity | Ovatools ≥0.514% | 69.1 (55.2; 80.9) | 92.4 (92.2; 92.7) |
| 40-49 years |  | CA125 >35 | 78.7 (72.7; 83.9) | 92.0 (91.9; 92.2) |
|  | sensitivity | Ovatools ≥0.51% | 78.7 (72.7; 83.9) | 91.3 (91.1; 91.5) |
|  | specificity | Ovatools >0.54% | 78.3 (72.3; 83.5) | 92.0 (91.8; 92.2) |
| <50 years |  | CA125 >35 | 75.3 (70.0; 80.0) | 92.5 (92.3; 92.6) |
|  | sensitivity | Ovatools ≥0.52% | 75.3 (70.0; 80.0) | 91.6 (91.5; 91.7) |
|  | specificity | Ovatools ≥0.57% | 73.6 (68.2; 78.5) | 92.5 (92.4; 92.7) |
| >50 years |  | CA125 >35 | 86.5 (84.8; 88.0) | 94.3 (94.2; 94.4) |
|  | sensitivity | Ovatools ≥2.12% | 86.5 (84.8; 88.0) | 94.8 (94.7; 94.9) |
|  | specificity | Ovatools ≥1.95% | 87.7 (86.1; 89.1) | 94.3 (94.2; 94.4) |

### Supplementary Table S1: Disease code list

| **Disease** | **Code** |
| --- | --- |
| Cancer |  |
| Ovarian | C56, C57.0, C48.1, C48.2 |
| Uterine | C54, C55 |
| Pancreatic | C25 |
| Upper gastrointestinal | C25, C22, C23, C24, C15, C16, C17 |
| Lower gastrointestinal | C18, C19, C20, C21 |
| All cancers | All ICD-10 C category except C44 (non-melanoma skin cancers) |
| Benign gynaecological disease | N70 – N77, N80 – N99, D10 – D48 |

### Supplementary Table S2: Summary of parameters in one-way sensitivity and scenario analyses

|  | **Base case** | **Scenarios** |
| --- | --- | --- |
| Relative risk ratio of late-stage diagnosis incidence | 0.836 | 0.737 and 0.950 |
| USS sensitivity | 0.85 | 0.80 and 0.90 |
| USS specificity | 0.83 | 0.78 and 0.88 |
| USS cost | 204 | 64 and 210 |
| One-year QoL utility adjustment due to surgery for benign disease | -0.04 | 0 |
| Annual QoL utility adjustment due to surgery for benign disease | 0.008 | 0 |
| Surgery rate for referred non-cancer cases | 0.75 | 0.60 and 0.90 |
| Annual QoL utility adjustment for late stage cancer in the long-term QoL model | -0.046 | -0.06 and 0 |
| Additional pathway effect on other cancers | See supplementary Method 9 | |
| Alternative CA125 and Ovatools accuracy | See supplementary Method 10 | |
| Duration of cancer-specific mortality prediction using the estimated survival model based on CPRD and linked data. | First 8 years | First 15 years |
| Annual discount rate | 3.5% | 1.5% |

### Supplementary Table S3: Characteristics of study population

|  | **Population with records of**  **CA125 or USS** | **Population with CA125 records** | **Population without CA125 but with USS records** |
| --- | --- | --- | --- |
|  | **Mean (SD) / n (%)** | | |
| N | 416,004 | 276,827 | 139,177 |
| Age | 50.6 (17.2) | 54.6 (15.8) | 42.7 (17.0) |
| Ethnicity |  |  |  |
| White | 359,698 (86.5) | 248,957 (89.9) | 110,741 (79.6) |
| Asian | 30,585 (7.4) | 15693 (5.7) | 14,892 (10.7) |
| Black | 20,170 (4.9) | 9379 (3.4) | 10,,791 ( 7.8) |
| Mixed or others | 5,551 (1.3) | 2,798 (1.0) | 2,753 ( 2.0) |
| Townsend Score |  |  |  |
| Q1 (least deprived) | 88,827 (21.4) | 67437 (24.4) | 21,390 (15.4) |
| Q2 | 83,582 (20.1) | 60760 (21.9) | 22,822 (16.4) |
| Q3 | 78,130 (18.8) | 53900 (19.5) | 24,230 (17.4) |
| Q4 | 74,474 (17.9) | 46959 (17.0) | 27,515 (19.8) |
| Q5 | 90,991 (21.9) | 47771 (17.3) | 43,220 (31.1) |
| Previous non-ovarian cancer | 27,335 (6.6) | 20,624 (7.5) | 6,711 ( 4.8) |
| Incident cancer diagnosed within 1 year |  |  |  |
| Invasive Ovarian cancer | 2,218 (0.53) | 1964 (0.71) | 254 (0.2) |
| Stage 1/2 | 515 (23.2) | 444 (22.6) | 71 (28.0) |
| Stage 3/4 | 1,056 (47.6) | 965 (49.1) | 91 (35.8) |
| Stage missing | 647 (29.2) | 555 (28.3) | 92 (36.2) |
| Lower GI cancer | 1,583 (0.38) | 1325 (0.48) | 258 (0.2) |
| Stage 1/2 | 498 (31.5) | 420 (31.7) | 78 (30.2) |
| Stage 3/4 | 864 (54.6) | 719 (54.3) | 145 (56.2) |
| Stage missing | 221 (14.0) | 186 (14.0) | 35 (13.6) |
| Uterine cancer | 1,049 (0.25) | 691 (0.25) | 358 (0.3) |
| Stage 1/2 | 619 (59.0) | 407 (58.9) | 212 (59.2) |
| Stage 3/4 | 238 (22.7) | 165 (23.9) | 73 (20.4) |
| Stage missing | 192 (18.3) | 119 (17.2) | 73 (20.4) |
| Lung cancer | 709 (0.17) | 593 (0.21) | 116 (0.1) |
| Stage 1/2 | 150 (21.2) | 125 (21.1) | 25 (21.6) |
| Stage 3/4 | 468 (66.0) | 394 (66.4) | 74 (63.8) |
| Stage missing | 91 (12.8) | 74 (12.5) | 17 (14.7) |
| Pancreatic cancer | 523 (0.13) | 441 (0.16) | 82 (0.1) |
| Stage 1/2 | 55 (10.5) | 46 (10.4) | 9 (11.0) |
| Stage 3/4 | 315 (60.2) | 264 (59.9) | 51 (62.2) |
| Stage missing | 153 (29.3) | 131 (29.7) | 22 (26.8) |
| Other cancers | 3,500 (0.84) | 2820 (1.0) | 680 (0.5) |
| Stage 1/2 | 1235 (35.3) | 955 (33.9) | 280 (41.2) |
| Stage 3/4 | 819 (23.4) | 677 (24.0) | 142 (20.9) |
| Stage missing | 1446 (41.3) | 1188 (42.1) | 258 (37.9) |
| Duration of follow up (years) | 5.46 (1.56) | 5.40 (1.58) | 5.58 (1.52) |

### Supplementary Table S4: Other pathway outcomes, costs and cost-effectiveness for women with CA125 records, compared with Pathway 1 (current practice)

|  | Proportion of stage shift* | ICER (£/LY gained) | LYs gained per 1000** | QALYs gained  per  1000** | Total cost difference per 1000 (£)** | Primary care pathway cost difference per 1000 (£) | FP referral related cost difference per 1000 (£) | Long term cost difference per 1000 (£)** | Long term cost difference per 1000 (£), discounted |
| --- | --- | --- | --- | --- | --- | --- | --- | --- | --- |
| Pathway 2 | | | | | | | | | |
| Age <50 | -0.021 | [-220,355] | -0.32 | -2.22 | -33260 | -11588 | -21986 | 314 | 221 |
| Age >=50 | 0.027 | 25489 | 2.07 | 2.34 | 35784 | 3237 | 30389 | 2157 | 1268 |
| Pathway 3 | | | | | | | | | |
| Age <50 | -0.016 | [-290,705] | -0.24 | -2.17 | -33384 | -11769 | -21854 | 239 | 168 |
| Age >=50 | 0.027 | 28956 | 2.05 | 2.44 | 40209 | 4633 | 33436 | 2140 | 1258 |
| Pathway 4 | | | | | | | | | |
| Age <50 | 0.045 | 775072 | 0.68 | 0.62 | 257960 | 193494 | 64997 | -531 | -408 |
| Age >=50 | 0.039 | 141508 | 3.02 | 2.87 | 284525 | 198937 | 82435 | 3154 | 1854 |
| Pathway 5 | | | | | | | | | |
| Age <50 | 0.045 | 785320 | 0.68 | 0.67 | 258973 | 193516 | 65983 | -526 | -404 |
| Age >=50 | 0.039 | 141773 | 3.01 | 2.87 | 284521 | 198938 | 82435 | 3148 | 1850 |
| Pathway 6 | | | | | | | | | |
| Age <50 | 0.053 | 864345 | 0.79 | 5.57 | 334452 | 195468 | 139601 | -617 | -475 |
| Age >=50 | 0.040 | 149138 | 3.08 | 3.55 | 306184 | 199384 | 103579 | 3221 | 1893 |

*Negative signs mean shift from early stage to late stage; **undiscounted. ICER, incremental cost-effectiveness ratio; LY, life year; QALY, quality adjusted life year; FP, false positive. ICERs in square brackets indicate £ saved per 1 less LY, due to reduction in benefit and cost saving compared to Pathway 1.

### Supplementary Table S5: Outcomes, costs and cost-effectiveness, by primary care pathway among women with CA125 or USS records

| **Pathway** | **Invasive OC detection rate (%)** | **Referral rate, among all patients (%)** | **QALYs gained per 1000** | **Cost difference per 1000 (£)** | **ICER (£/QALY gained)**** |
| --- | --- | --- | --- | --- | --- |
| **Age ≤ 49 years:** N = 210,552; 285 (0.14%) invasive OC with 44% late stage | | | | |  |
| **Pathway 1 (current practice)** | 64 | 1.4 |  |  |  |
| Pathway 2 | 59 | 1.1 | -0.95 | -33,327 | [-34,999] |
| Pathway 3 | 60 | 1.2 | -0.94 | -33,407 | [-35,616] |
| Pathway 4 | 92 | 18 | 0.23 | 259,095 | ED |
| Pathway 5 | 92 | 18 | 0.25 | 260,123 | ED |
| Pathway 6 | 96 | 23 | 2.47 | 337,268 | 136,726 |
| **Age ≥ 50 years:** N = 205,452; 1933 (0.94%) invasive OC with 71% late-stage | | | | |  |
| **Pathway 1 (current practice)** | 74 | 1.7 |  |  |  |
| Pathway 2 | 90 | 5.6 | 1.36 | 35,295 | 26,000 |
| Pathway 3 | 90 | 5.7 | 1.41 | 39,735 | 28,175 |
| Pathway 4 | 97 | 21 | 1.68 | 283,682 | ED |
| Pathway 5 | 97 | 21 | 1.67 | 283,680 | ED |
| Pathway 6 | 98 | 22 | 2.05 | 305,396 | 392,360 |

*Compared with Pathway 1; QALYs and cost were discounted. **Compared with the non-dominated less costly alternative (Ovatools and its equivalent age-adjusted threshold pathways were not compared with each other). ICERs in square brackets indicate £ saved per 1 less QALY, due to reduction in benefit and cost saving compared to Pathway 1.

CA125, cancer antigen 125; OC, ovarian cancer; QALY, Quality-adjusted life year; ICER, incremental cost-effectiveness ratio; USS, ultrasound scan; ED, extendedly dominated.

### Supplementary Table S6: Other pathway outcomes, costs and cost-effectiveness for women with CA125 or USS records, compared with Pathway 1 (current practice)

|  | Proportion of stage shift* | ICER (£/LY gained) | LYs gained per 1000** | QALYs gained per 1000** | Total cost difference per 1000 (£)** | Primary care pathway cost difference per 1000 (£) | FP referral related cost difference per 1000 (£) | Long term cost difference per 1000 (£)** | Long term cost difference per 1000 (£), discounted |
| --- | --- | --- | --- | --- | --- | --- | --- | --- | --- |
| Pathway 2 | | | | | | | | | |
| Age <50 | -0.022 | [-331,298] | -0.21 | -2.26 | -33240 | -11498 | -21995 | 253 | 167 |
| Age >=50 | 0.027 | 29182 | 1.83 | 2.17 | 36086 | 3458 | 30717 | 1911 | 1120 |
| Pathway 3 | | | | | | | | | |
| Age <50 | -0.017 | [-436,797] | -0.16 | -2.23 | -33342 | -11683 | -21851 | 192 | 127 |
| Age >=50 | 0.027 | 33116 | 1.81 | 2.27 | 40520 | 4855 | 33769 | 1896 | 1112 |
| Pathway 4 | | | | | | | | | |
| Age <50 | 0.045 | 1212980 | 0.44 | 0.49 | 258984 | 193526 | 65860 | -401 | -290 |
| Age >=50 | 0.039 | 160431 | 2.67 | 2.61 | 284838 | 199169 | 82875 | 2794 | 1638 |
| Pathway 5 | | | | | | | | | |
| Age <50 | 0.045 | 1229075 | 0.44 | 0.55 | 260013 | 193549 | 66862 | -398 | -288 |
| Age >=50 | 0.039 | 160732 | 2.67 | 2.60 | 284834 | 199170 | 82875 | 2789 | 1635 |
| Pathway 6 | | | | | | | | | |
| Age <50 | 0.053 | 1358174 | 0.51 | 5.85 | 337139 | 195620 | 141986 | -467 | -337 |
| Age >=50 | 0.040 | 169108 | 2.73 | 3.28 | 306576 | 199621 | 104102 | 2854 | 1673 |

*Negative signs mean shift from early stage to late stage; **undiscounted. ICER, incremental cost-effectiveness ratio; LY, life year; QALY, quality adjusted life year; FP, false positive. ICERs in square brackets indicate £ saved per 1 less LY, due to reduction in benefit and cost saving compared to the current pathway.

### Supplementary Table S7: Cost-effectiveness data of the Ovatools Sequential pathway, compared Pathway 1, using different Ovatools risk thresholds for women aged ≥ 50 years with CA125 records (N = 164,746)

| Moderate risk threshold (%) | High risk threshold (%) | ICER (£/QALY gained) | ICER (£/LY gained) | Total Life year gained* | Total QALYs gained* | Additional long-term inpatient care cost* | Additional primary care pathway cost* | Additional cost related to false positive referral* |
| --- | --- | --- | --- | --- | --- | --- | --- | --- |
| 0.6 | 2 | 33550 | 43469 | 384 | 536 | 400784 | 1561127 | 9258621 |
| 0.6 | 2.2 | 33623 | 44186 | 379 | 538 | 396226 | 1760261 | 9116812 |
| 0.6 | 2.4 | 33637 | 44697 | 377 | 541 | 393296 | 1925219 | 8998951 |
| 0.6 | 2.6 | 33663 | 45152 | 374 | 543 | 390691 | 2057633 | 8906851 |
| 0.6 | 2.8 | 33701 | 45542 | 372 | 544 | 388412 | 2157428 | 8839238 |
| 0.6 | 3 | 33704 | 45858 | 370 | 546 | 386784 | 2256438 | 8771637 |
| 0.6 | 3.2 | 33828 | 46457 | 367 | 546 | 382877 | 2358341 | 8704057 |
| 0.6 | 3.4 | 33929 | 46910 | 364 | 546 | 379947 | 2426734 | 8660058 |
| 0.6 | 3.6 | 34013 | 47333 | 361 | 546 | 377342 | 2494774 | 8617466 |
| 0.6 | 3.8 | 34073 | 47593 | 360 | 546 | 375715 | 2529234 | 8597037 |
| 0.6 | 4 | 34158 | 48036 | 357 | 547 | 373110 | 2597357 | 8556519 |
| 0.8 | 2 | 28329 | 32306 | 368 | 445 | 384179 | 566931 | 7083096 |
| 0.8 | 2.2 | 28431 | 32919 | 363 | 447 | 379621 | 765783 | 6941287 |
| 0.8 | 2.4 | 28470 | 33364 | 361 | 450 | 376691 | 930511 | 6823468 |
| 0.8 | 2.6 | 28516 | 33760 | 358 | 452 | 374087 | 1062736 | 6731431 |
| 0.8 | 2.8 | 28570 | 34097 | 356 | 453 | 371808 | 1162384 | 6663817 |
| 0.8 | 3 | 28589 | 34376 | 354 | 455 | 370180 | 1261250 | 6596217 |
| 0.8 | 3.2 | 28730 | 34879 | 351 | 455 | 366273 | 1362998 | 6528636 |
| 0.8 | 3.4 | 28843 | 35259 | 348 | 455 | 363343 | 1431285 | 6484638 |
| 0.8 | 3.6 | 28940 | 35617 | 345 | 455 | 360738 | 1499217 | 6442045 |
| 0.8 | 3.8 | 29007 | 35836 | 344 | 455 | 359110 | 1533620 | 6421616 |
| 0.8 | 4 | 29106 | 36212 | 341 | 455 | 356506 | 1601632 | 6381098 |
| 1 | 2 | 23218 | 23676 | 354 | 376 | 369420 | -160268 | 5493290 |
| 1 | 2.2 | 23356 | 24205 | 349 | 378 | 364862 | 38381 | 5351480 |
| 1 | 2.4 | 23425 | 24598 | 347 | 381 | 361932 | 202942 | 5233692 |
| 1 | 2.6 | 23498 | 24945 | 344 | 383 | 359327 | 335031 | 5141700 |
| 1 | 2.8 | 23571 | 25239 | 342 | 384 | 357048 | 434573 | 5074087 |
| 1 | 3 | 23610 | 25489 | 340 | 386 | 355420 | 533334 | 5006486 |
| 1 | 3.2 | 23768 | 25914 | 337 | 386 | 351513 | 634971 | 4938905 |
| 1 | 3.4 | 23892 | 26234 | 334 | 386 | 348583 | 703180 | 4894907 |
| 1 | 3.6 | 24002 | 26539 | 331 | 386 | 345979 | 771035 | 4852314 |
| 1 | 3.8 | 24075 | 26724 | 330 | 386 | 344351 | 805396 | 4831886 |
| 1 | 4 | 24188 | 27047 | 327 | 386 | 341746 | 873327 | 4791367 |
| 1.2 | 2 | 17736 | 16257 | 345 | 323 | 360195 | -734028 | 4238179 |
| 1.2 | 2.2 | 17915 | 16705 | 340 | 325 | 355637 | -535542 | 4096369 |
| 1.2 | 2.4 | 18025 | 17044 | 338 | 328 | 352707 | -371113 | 3978606 |
| 1.2 | 2.6 | 18130 | 17344 | 335 | 330 | 350103 | -239134 | 3886650 |
| 1.2 | 2.8 | 18227 | 17596 | 333 | 331 | 347824 | -139677 | 3819036 |
| 1.2 | 3 | 18293 | 17816 | 331 | 333 | 346196 | -40999 | 3751436 |
| 1.2 | 3.2 | 18465 | 18165 | 328 | 333 | 342289 | 60548 | 3683855 |
| 1.2 | 3.4 | 18598 | 18426 | 325 | 333 | 339359 | 128696 | 3639857 |
| 1.2 | 3.6 | 18720 | 18679 | 322 | 333 | 336754 | 196488 | 3597264 |
| 1.2 | 3.8 | 18798 | 18831 | 321 | 333 | 335126 | 230817 | 3576835 |
| 1.2 | 4 | 18925 | 19101 | 318 | 334 | 332521 | 298683 | 3536317 |
| 1.4 | 2 | 11804 | 9720 | 329 | 271 | 343591 | -1233025 | 3150416 |
| 1.4 | 2.2 | 12043 | 10101 | 325 | 273 | 339033 | -1034672 | 3008607 |
| 1.4 | 2.4 | 12210 | 10400 | 322 | 276 | 336103 | -870353 | 2890877 |
| 1.4 | 2.6 | 12360 | 10662 | 319 | 278 | 333498 | -738464 | 2798939 |
| 1.4 | 2.8 | 12490 | 10881 | 317 | 279 | 331219 | -639077 | 2731326 |
| 1.4 | 3 | 12593 | 11078 | 316 | 281 | 329591 | -540467 | 2663725 |
| 1.4 | 3.2 | 12785 | 11364 | 312 | 281 | 325684 | -438993 | 2596144 |
| 1.4 | 3.4 | 12930 | 11576 | 309 | 281 | 322754 | -370896 | 2552146 |
| 1.4 | 3.6 | 13067 | 11787 | 307 | 281 | 320150 | -303156 | 2509554 |
| 1.4 | 3.8 | 13152 | 11911 | 305 | 281 | 318522 | -268854 | 2489125 |
| 1.4 | 4 | 13297 | 12138 | 302 | 281 | 315917 | -201041 | 2448607 |
| 1.6 | 2 | 5672 | 4230 | 318 | 232 | 332521 | -1616611 | 2313676 |
| 1.6 | 2.2 | 5979 | 4547 | 314 | 235 | 327963 | -1418363 | 2171867 |
| 1.6 | 2.4 | 6215 | 4805 | 311 | 237 | 325033 | -1254128 | 2054163 |
| 1.6 | 2.6 | 6419 | 5031 | 309 | 239 | 322429 | -1122310 | 1962239 |
| 1.6 | 2.8 | 6586 | 5217 | 307 | 241 | 320150 | -1022977 | 1894626 |
| 1.6 | 3 | 6736 | 5392 | 305 | 242 | 318522 | -924421 | 1827025 |
| 1.6 | 3.2 | 6943 | 5617 | 301 | 242 | 314615 | -823005 | 1759444 |
| 1.6 | 3.4 | 7096 | 5782 | 298 | 242 | 311685 | -754947 | 1715446 |
| 1.6 | 3.6 | 7247 | 5951 | 296 | 243 | 309080 | -687247 | 1672853 |
| 1.6 | 3.8 | 7336 | 6049 | 294 | 242 | 307452 | -652967 | 1652425 |
| 1.6 | 4 | 7499 | 6234 | 292 | 243 | 304848 | -585195 | 1611906 |
| 1.8 | 2 | -701 | -474 | 302 | 195 | 315917 | -1925046 | 1644284 |
| 1.8 | 2.2 | -303 | -210 | 298 | 198 | 311359 | -1726874 | 1502474 |
| 1.8 | 2.4 | 24 | 17 | 295 | 200 | 308429 | -1562703 | 1384791 |
| 1.8 | 2.6 | 301 | 215 | 293 | 202 | 305824 | -1430936 | 1292879 |
| 1.8 | 2.8 | 517 | 374 | 291 | 204 | 303545 | -1331644 | 1225265 |
| 1.8 | 3 | 728 | 532 | 289 | 205 | 301917 | -1233127 | 1157665 |
| 1.8 | 3.2 | 955 | 706 | 285 | 206 | 298010 | -1131754 | 1090084 |
| 1.8 | 3.4 | 1117 | 832 | 283 | 205 | 295080 | -1063726 | 1046086 |
| 1.8 | 3.6 | 1287 | 966 | 280 | 206 | 292476 | -996055 | 1003493 |
| 1.8 | 3.8 | 1381 | 1041 | 278 | 205 | 290848 | -961791 | 983064 |
| 1.8 | 4 | 1567 | 1191 | 276 | 206 | 288243 | -894051 | 942546 |
| 2 | 2.2 | -7713 | -4813 | 284 | 165 | 296600 | -1996918 | 916756 |
| 2 | 2.4 | -7257 | -4621 | 281 | 168 | 293669 | -1832801 | 799091 |
| 2 | 2.6 | -6882 | -4455 | 279 | 170 | 291065 | -1701080 | 707188 |
| 2 | 2.8 | -6598 | -4325 | 276 | 171 | 288786 | -1601823 | 639575 |
| 2 | 3 | -6303 | -4185 | 275 | 173 | 287158 | -1503340 | 571974 |
| 2 | 3.2 | -6057 | -4068 | 271 | 173 | 283251 | -1402004 | 504394 |
| 2 | 3.4 | -5889 | -3985 | 268 | 173 | 280321 | -1334002 | 460395 |
| 2 | 3.6 | -5699 | -3889 | 266 | 173 | 277716 | -1266357 | 417803 |
| 2 | 3.8 | -5601 | -3838 | 264 | 173 | 276088 | -1232107 | 397374 |
| 2 | 4 | -5390 | -3727 | 262 | 173 | 273484 | -1164394 | 356856 |

*Total, undiscounted. ICER, incremental cost-effectiveness ratio; LY, life year; QALY, quality adjusted life year

### Supplementary Table S8: Cost-effectiveness data of the Ovatools Sequential pathway, compared with Pathway 1, using different Ovatools risk thresholds for women aged ≥ 50 years with CA125 or USS records (N=205,452)

| Moderate risk threshold | High risk threshold | ICER (£/QALY gained) | ICER (£/LY gained) | LYs gained* | QALYs gained* | Additional long-term inpatient care cost* | Additional primary care pathway cost* | Additional cost related to false positive referral* |
| --- | --- | --- | --- | --- | --- | --- | --- | --- |
| 0.6 | 2 | 36234 | 49587 | 423 | 629 | 442762 | 1995615 | 11639496 |
| 0.6 | 2.2 | 36268 | 50395 | 418 | 633 | 437726 | 2243468 | 11460510 |
| 0.6 | 2.4 | 36248 | 50969 | 415 | 636 | 434489 | 2448918 | 11311491 |
| 0.6 | 2.6 | 36247 | 51470 | 413 | 639 | 431612 | 2613718 | 11192348 |
| 0.6 | 2.8 | 36265 | 51895 | 410 | 641 | 429094 | 2737823 | 11103242 |
| 0.6 | 3 | 36247 | 52237 | 409 | 643 | 427296 | 2861058 | 11014252 |
| 0.6 | 3.2 | 36352 | 52904 | 404 | 643 | 422980 | 2987543 | 10926185 |
| 0.6 | 3.4 | 36440 | 53405 | 401 | 643 | 419743 | 3072363 | 10868094 |
| 0.6 | 3.6 | 36511 | 53871 | 399 | 644 | 416865 | 3156773 | 10810986 |
| 0.6 | 3.8 | 36564 | 54154 | 397 | 644 | 415067 | 3199440 | 10782432 |
| 0.6 | 4 | 36635 | 54630 | 394 | 644 | 412189 | 3283878 | 10725324 |
| 0.8 | 2 | 30988 | 36953 | 406 | 517 | 424418 | 754707 | 8921944 |
| 0.8 | 2.2 | 31051 | 37644 | 401 | 520 | 419383 | 1002210 | 8742971 |
| 0.8 | 2.4 | 31055 | 38142 | 398 | 523 | 416146 | 1207374 | 8593953 |
| 0.8 | 2.6 | 31072 | 38575 | 395 | 526 | 413268 | 1371941 | 8474809 |
| 0.8 | 2.8 | 31104 | 38940 | 393 | 528 | 410751 | 1495870 | 8385703 |
| 0.8 | 3 | 31100 | 39239 | 391 | 530 | 408952 | 1618933 | 8296713 |
| 0.8 | 3.2 | 31223 | 39797 | 387 | 530 | 404636 | 1745232 | 8208646 |
| 0.8 | 3.4 | 31324 | 40215 | 384 | 530 | 401399 | 1829924 | 8150556 |
| 0.8 | 3.6 | 31407 | 40606 | 381 | 531 | 398522 | 1914207 | 8093447 |
| 0.8 | 3.8 | 31467 | 40842 | 379 | 531 | 396723 | 1956809 | 8064893 |
| 0.8 | 4 | 31550 | 41243 | 377 | 531 | 393846 | 2041120 | 8007785 |
| 1 | 2 | 25755 | 27189 | 390 | 432 | 408113 | -152867 | 6936050 |
| 1 | 2.2 | 25856 | 27785 | 385 | 435 | 403077 | 94382 | 6757077 |
| 1 | 2.4 | 25892 | 28224 | 382 | 439 | 399840 | 299337 | 6608059 |
| 1 | 2.6 | 25935 | 28602 | 380 | 441 | 396963 | 463737 | 6488915 |
| 1 | 2.8 | 25985 | 28917 | 377 | 443 | 394445 | 587539 | 6399809 |
| 1 | 3 | 26000 | 29182 | 375 | 445 | 392647 | 710476 | 6310819 |
| 1 | 3.2 | 26141 | 29651 | 371 | 446 | 388331 | 836641 | 6222753 |
| 1 | 3.4 | 26254 | 30001 | 368 | 446 | 385094 | 921241 | 6164662 |
| 1 | 3.6 | 26350 | 30332 | 365 | 446 | 382216 | 1005431 | 6107554 |
| 1 | 3.8 | 26416 | 30529 | 364 | 446 | 380418 | 1047986 | 6079000 |
| 1 | 4 | 26512 | 30868 | 361 | 446 | 377540 | 1132205 | 6021891 |
| 1.2 | 2 | 20018 | 18793 | 380 | 367 | 397922 | -868996 | 5368239 |
| 1.2 | 2.2 | 20168 | 19295 | 376 | 370 | 392887 | -621949 | 5189267 |
| 1.2 | 2.4 | 20249 | 19674 | 373 | 374 | 389650 | -417161 | 5040248 |
| 1.2 | 2.6 | 20327 | 19998 | 370 | 377 | 386772 | -252894 | 4921105 |
| 1.2 | 2.8 | 20402 | 20265 | 367 | 378 | 384254 | -129194 | 4831998 |
| 1.2 | 3 | 20444 | 20496 | 366 | 381 | 382456 | -6357 | 4743008 |
| 1.2 | 3.2 | 20602 | 20878 | 362 | 381 | 378140 | 119700 | 4654942 |
| 1.2 | 3.4 | 20726 | 21162 | 358 | 381 | 374903 | 204227 | 4596851 |
| 1.2 | 3.6 | 20835 | 21433 | 356 | 381 | 372025 | 288344 | 4539743 |
| 1.2 | 3.8 | 20906 | 21592 | 354 | 381 | 370227 | 330861 | 4511189 |
| 1.2 | 4 | 21015 | 21870 | 351 | 382 | 367350 | 415006 | 4454080 |
| 1.4 | 2 | 13690 | 11408 | 363 | 304 | 379579 | -1491582 | 4009470 |
| 1.4 | 2.2 | 13913 | 11835 | 358 | 307 | 374543 | -1244702 | 3830497 |
| 1.4 | 2.4 | 14060 | 12167 | 355 | 311 | 371306 | -1040051 | 3681479 |
| 1.4 | 2.6 | 14188 | 12448 | 352 | 313 | 368429 | -875895 | 3562335 |
| 1.4 | 2.8 | 14298 | 12676 | 350 | 315 | 365911 | -752279 | 3473229 |
| 1.4 | 3 | 14379 | 12881 | 348 | 317 | 364113 | -629524 | 3384239 |
| 1.4 | 3.2 | 14561 | 13191 | 344 | 318 | 359796 | -503557 | 3296172 |
| 1.4 | 3.4 | 14698 | 13419 | 341 | 318 | 356559 | -419091 | 3238082 |
| 1.4 | 3.6 | 14825 | 13641 | 338 | 318 | 353682 | -335035 | 3180973 |
| 1.4 | 3.8 | 14902 | 13769 | 336 | 318 | 351884 | -292549 | 3152419 |
| 1.4 | 4 | 15029 | 13997 | 334 | 318 | 349006 | -208465 | 3095311 |
| 1.6 | 2 | 6981 | 5202 | 351 | 256 | 367350 | -1970210 | 2964263 |
| 1.6 | 2.2 | 7293 | 5557 | 346 | 260 | 362314 | -1723460 | 2785290 |
| 1.6 | 2.4 | 7527 | 5843 | 343 | 263 | 359077 | -1518916 | 2636271 |
| 1.6 | 2.6 | 7718 | 6082 | 341 | 266 | 356200 | -1354847 | 2517128 |
| 1.6 | 2.8 | 7871 | 6273 | 338 | 268 | 353682 | -1231296 | 2428022 |
| 1.6 | 3 | 8004 | 6452 | 336 | 270 | 351884 | -1108605 | 2339032 |
| 1.6 | 3.2 | 8207 | 6692 | 332 | 270 | 347567 | -982707 | 2250965 |
| 1.6 | 3.4 | 8356 | 6867 | 329 | 270 | 344330 | -898289 | 2192874 |
| 1.6 | 3.6 | 8499 | 7042 | 326 | 271 | 341453 | -814280 | 2135766 |
| 1.6 | 3.8 | 8581 | 7139 | 325 | 271 | 339655 | -771818 | 2107212 |
| 1.6 | 4 | 8725 | 7318 | 322 | 271 | 336777 | -687782 | 2050104 |
| 1.8 | 2 | -150 | -100 | 334 | 212 | 349006 | -2354858 | 2128097 |
| 1.8 | 2.2 | 286 | 196 | 329 | 215 | 343971 | -2108206 | 1949124 |
| 1.8 | 2.4 | 639 | 446 | 326 | 219 | 340734 | -1903741 | 1800106 |
| 1.8 | 2.6 | 918 | 652 | 323 | 222 | 337856 | -1739736 | 1680962 |
| 1.8 | 2.8 | 1129 | 812 | 321 | 223 | 335338 | -1616234 | 1591856 |
| 1.8 | 3 | 1333 | 970 | 319 | 225 | 333540 | -1493591 | 1502866 |
| 1.8 | 3.2 | 1564 | 1152 | 315 | 226 | 329224 | -1367745 | 1414799 |
| 1.8 | 3.4 | 1726 | 1282 | 312 | 226 | 325987 | -1283362 | 1356709 |
| 1.8 | 3.6 | 1891 | 1416 | 309 | 226 | 323109 | -1199388 | 1299600 |
| 1.8 | 3.8 | 1978 | 1488 | 307 | 226 | 321311 | -1156945 | 1271046 |
| 1.8 | 4 | 2143 | 1626 | 304 | 226 | 318434 | -1072944 | 1213938 |
| 2 | 2.2 | -8214 | -4993 | 313 | 176 | 327665 | -2444966 | 1217479 |
| 2 | 2.4 | -7685 | -4782 | 310 | 180 | 324428 | -2240571 | 1068461 |
| 2 | 2.6 | -7278 | -4613 | 307 | 183 | 321551 | -2076621 | 949317 |
| 2 | 2.8 | -6983 | -4487 | 305 | 184 | 319033 | -1953162 | 860211 |
| 2 | 3 | -6675 | -4350 | 303 | 186 | 317235 | -1830561 | 771221 |
| 2 | 3.2 | -6413 | -4232 | 299 | 187 | 312919 | -1704760 | 683154 |
| 2 | 3.4 | -6239 | -4152 | 296 | 187 | 309681 | -1620407 | 625064 |
| 2 | 3.6 | -6049 | -4062 | 293 | 187 | 306804 | -1536465 | 567955 |
| 2 | 3.8 | -5960 | -4019 | 292 | 187 | 305006 | -1494037 | 539401 |
| 2 | 4 | -5769 | -3926 | 289 | 187 | 302128 | -1410067 | 482293 |

*Total, undiscounted. ICER, incremental cost-effectiveness ratio; LY, life year; QALY, quality adjusted life year

### Supplementary Table S9: Age-group specific CA125 thresholds corresponding to the Ovatools risk levels

|  | Matched CA125 Thresholds (U/mL) | | | | | | |
| --- | --- | --- | --- | --- | --- | --- | --- |
|  | 18-29 years | 30-39 years | 40-49 years | 50-59 years | 60-69 years | 70-79 years | 80-89 years |
| Ovatools ≥0.6% | 26 | 38 | 37 | 22 | 19 | 20 | 22 |
| Ovatools ≥0.8% | 30 | 49 | 48 | 24 | 20 | 21 | 24 |
| Ovatools ≥1% | 34 | 59 | 58 | 26 | 22 | 22 | 26 |
| Ovatools ≥1.2% | 40 | 70 | 68 | 28 | 23 | 24 | 28 |
| Ovatools ≥1.4% | 45 | 80 | 79 | 31 | 24 | 25 | 31 |
| Ovatools ≥1.6% | 51 | 91 | 89 | 33 | 25 | 27 | 33 |
| Ovatools ≥1.8% | 57 | 101 | 99 | 36 | 27 | 28 | 36 |
| Ovatools ≥2% | 63 | 111 | 108 | 40 | 28 | 30 | 40 |
| Ovatools ≥2.2% | 68 | 121 | 118 | 43 | 30 | 32 | 44 |
| Ovatools ≥2.4% | 74 | 131 | 128 | 47 | 31 | 34 | 47 |
| Ovatools ≥2.6% | 79 | 140 | 137 | 50 | 33 | 36 | 51 |
| Ovatools ≥2.8% | 85 | 150 | 147 | 54 | 35 | 39 | 54 |
| Ovatools ≥3% | 91 | 160 | 157 | 57 | 37 | 41 | 58 |
| Ovatools ≥3.2% | 96 | 170 | 166 | 61 | 39 | 43 | 61 |
| Ovatools ≥3.4% | 102 | 179 | 176 | 64 | 41 | 46 | 65 |
| Ovatools ≥3.6% | 107 | 189 | 185 | 68 | 43 | 48 | 68 |
| Ovatools ≥3.8% | 112 | 199 | 195 | 71 | 45 | 51 | 72 |
| Ovatools ≥4% | 118 | 208 | 204 | 75 | 48 | 53 | 75 |

### Supplementary Table S10: Key study assumptions

|  | **Assumption** |
| --- | --- |
| Study population | Women in CPRD with CA125 records or USS records and related OC symptoms represent OC suspected patients in UK primary care. |
|  | All patients with cancer are diagnosed within one year of testing. |
| Resource use | Each patient presenting OC related symptoms receive an initial face-to-face GP consultation, followed by a CA125 test and/or USS, depending on the pathway. In sequential pathways, if a patient’s CA125 test result indicated a further USS investigation, she received a GP telephone follow-up for informing the result and USS appointment. A further GP consultation occurs along with referral to secondary care. The CA125 test requires 10-minute nurse time. GP face-to-face consultation requires 10 minutes and telephone follow-up requires 5 minutes. |
|  | For false negative cases in primary care OC detection, we assume the diagnostic process in primary care would be repeated once, so the primary care pathway cost is doubled. |
|  | After referral to hospital, all patients receive an outpatient consultation, a CA125 test and a USS. In the scenario of including effects on other cancers, the CT cost in secondary care for detecting other cancers is also included. |
|  | Costs for cancer inpatient treatment and surgery for benign gynaecological diseases are informed from NHS HRG reference costs. |
| Reduction of late-stage diagnosis incidence | Symptomatic women in the UKCTOCS are similar to symptomatic women seen in primary care |
|  | A fraction of additionally detected ‘previously late stage’ cancers ‘shift’ to an early stage at diagnosis in the new pathway, informed from findings from screening of symptomatic women in UKCTOCS. |
|  | Late-to-early shifted cancer cases have the same risk of cancer death as that of an early-stage cancer diagnosis |
| Decision tree | CA125/Ovatools and USS accuracies are independent |
|  | In the concurrent pathway, either exceeding the CA125/Ovatools threshold or having an abnormal USS finding leads to a referral |
| Markov model | Patients survive with cancer over 8 years have the same cancer-specific mortality rate (death caused by cancer) as the general population in the same age group, with the assumption extended to 15 years in the sensitivity analysis |
|  | Non-cancer mortality (death not caused by cancer) rate by age and sex is the same for people with and without cancer |
| Quality of life | The surgery for benign gynaecological disease reduces quality of life utility by 0.04 in the year of the surgery |
|  | The surgery for benign gynaecological disease increases quality of life utility by 0.008 annually after the surgery |
|  | Late-stage cancer diagnosis further reduce quality of life utility by 0.046 |

### Supplementary Figure S1: The Markov model structure for long term extrapolation

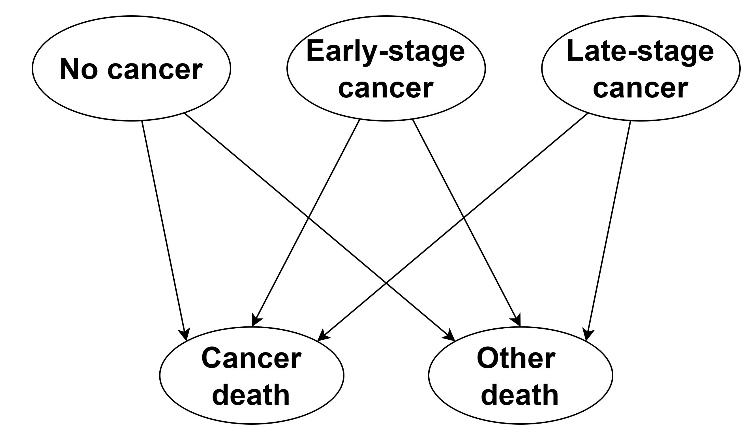

This structure applies to all cancer types and non-cancer as individuals enter from one of the three entering state.

### Supplementary Figure S2: Probabilistic outcome and sensitivity analyses for women aged ≥ 50 years with CA125 or USS records

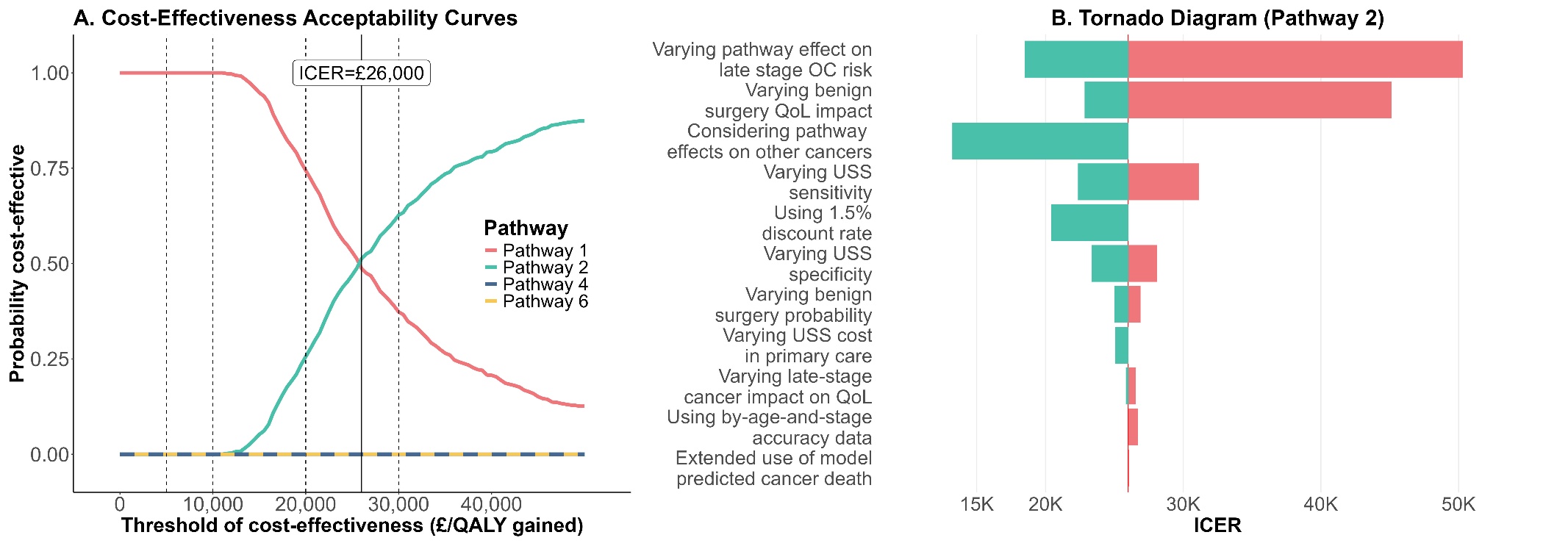

Panel B: See supplementary Table S2 for the base case values and ranges used in the sensitivity or scenario analyses

### Supplementary Figure S3: Cost-Effectiveness Acceptability Curve for women aged ≥ 50 years using probabilistic sensitivity analysis data, for Ovatools equivalent age-adjusted threshold pathways

A. **Among the women with CA125 records**

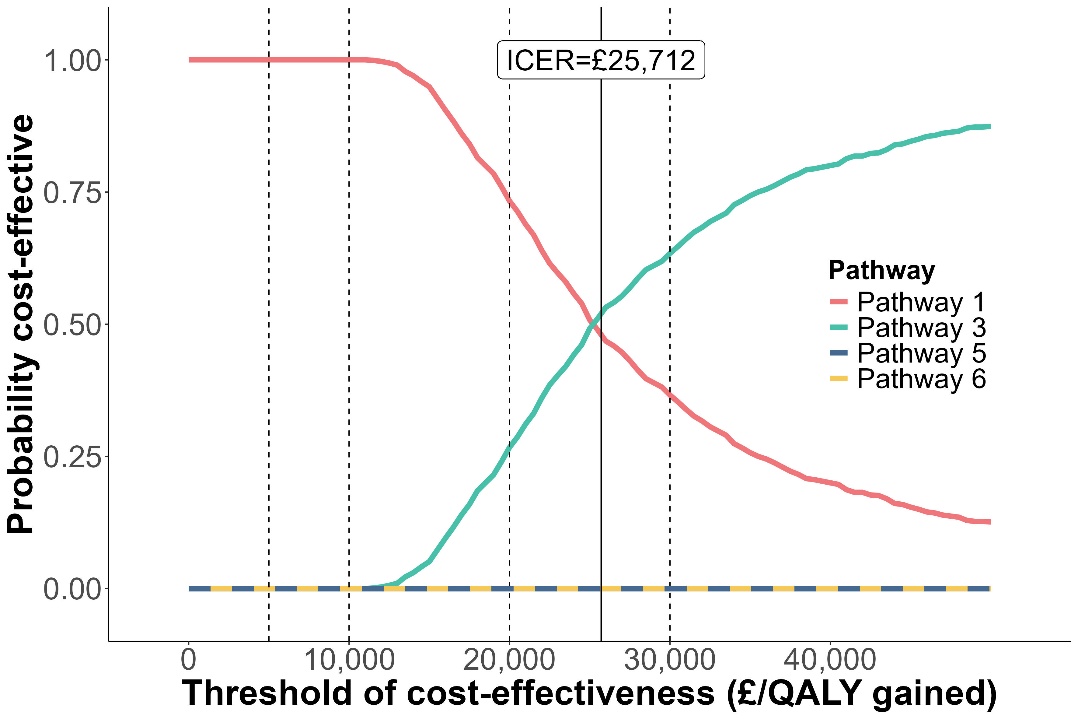

B. **Among the women with CA125 or USS records**

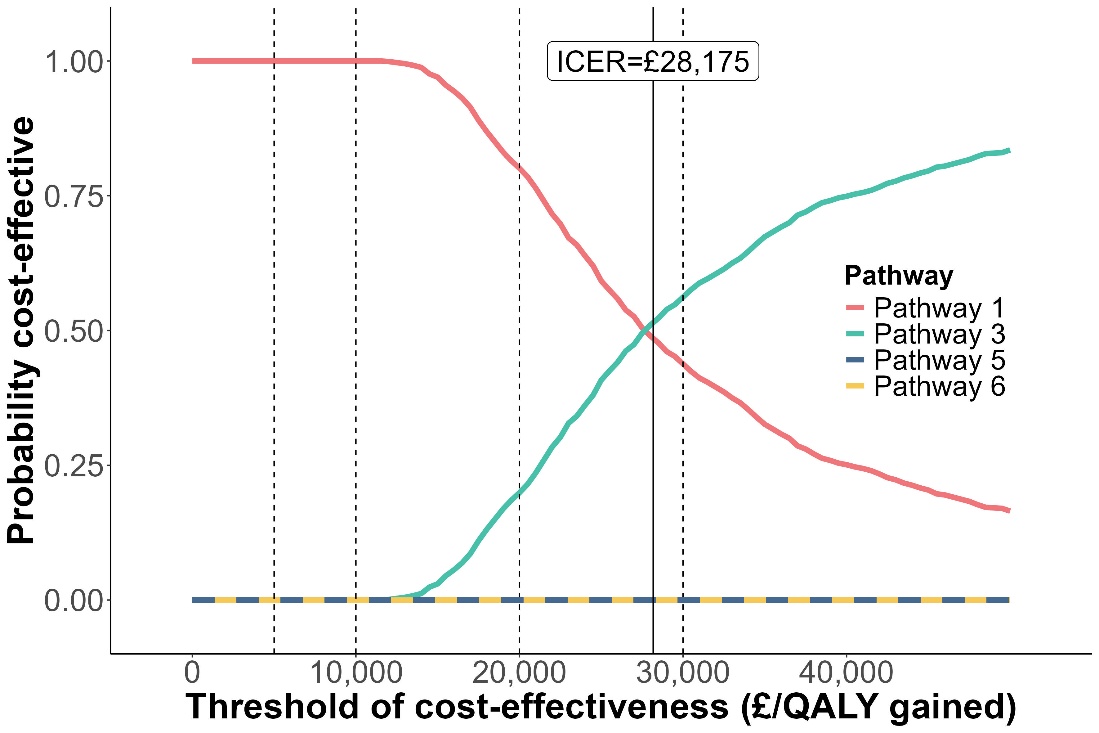

### Supplementary Figure S4: The heatmap of ICERs of Pathway 2 compared with Pathway 1 with different risk thresholds for moderate and high risks, among women with CA125 or USS records

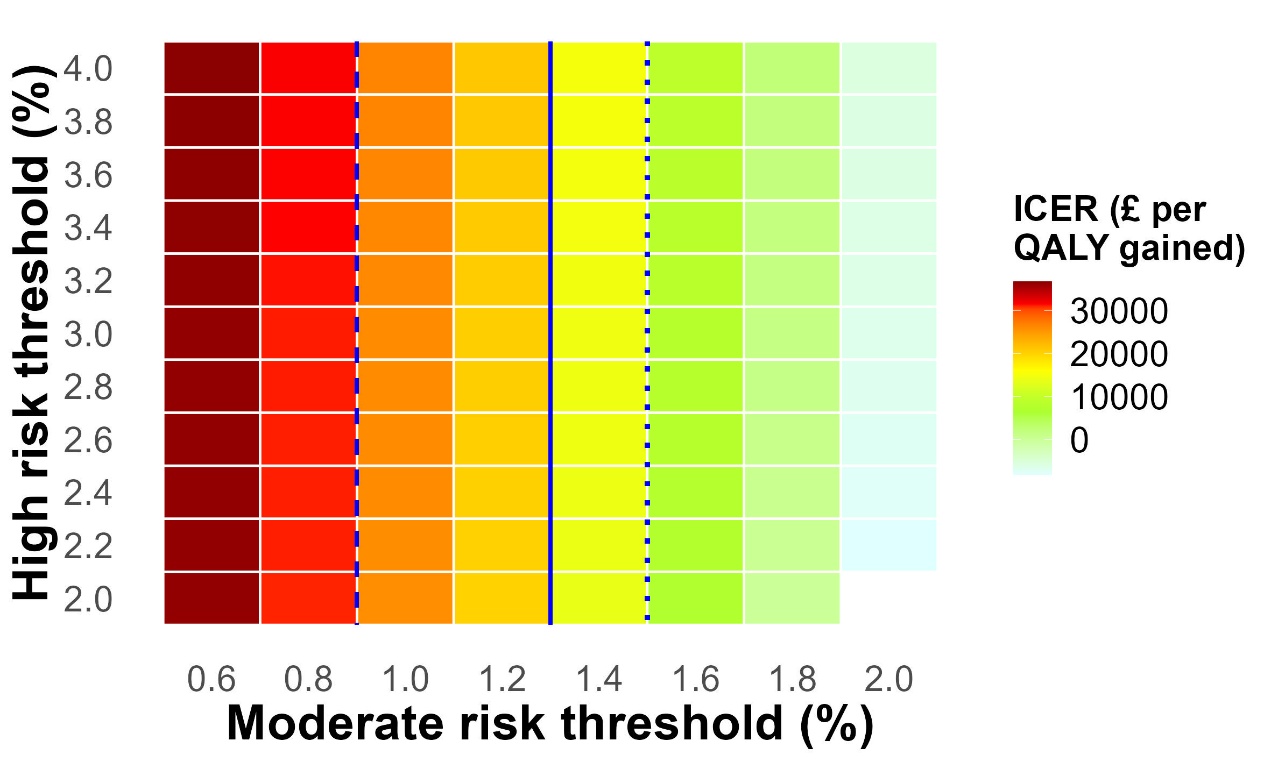

9. NHS England. Cancer Survival in England, cancers diagnosed 2016 to 2020, 2023.

10. NHS England. Costing in the NHS [Available from: <https://www.england.nhs.uk/costing-in-the-nhs/> accessed 17/11/2023.

11. NHS England. NHS Payment Scheme [Available from: <https://www.england.nhs.uk/pay-syst/nhs-payment-scheme/> accessed 17/11/2023.

12. Jones KC, Weatherly H, Birch S, et al. Unit costs of health and social care 2022 manual. 2022

13. Jones KC, Burns A. Unit costs of health and social care 2021. 2021

14. Zhou J, Williams C, Keng MJ, et al. Estimating Costs Associated with Disease Model States Using Generalized Linear Models: A Tutorial. *PharmacoEconomics* 2023:1-13.

15. Zhou J, Wu R, Williams C, et al. Prediction Models for Individual-Level Healthcare Costs Associated with Cardiovascular Events in the UK. *PharmacoEconomics* 2023;41(5):547-59.

16. Sudlow C, Gallacher J, Allen N, et al. UK biobank: an open access resource for identifying the causes of a wide range of complex diseases of middle and old age. *PLoS medicine* 2015;12(3):e1001779.

17. Hernandez Alava M, Pudney S, Wailoo A. Estimating the relationship between EQ-5D-5L and EQ-5D-3L: results from a UK population study. *Pharmacoeconomics* 2023;41(2):199-207. doi: <https://doi.or/10.1007/s40273-022-01218-7> [published Online First: 2022/12/01]

18. NHS England. Cancer Quality of Life Survey, 2021.

19. Funston G, Hamilton W, Abel G, et al. The diagnostic performance of CA125 for the detection of ovarian and non-ovarian cancer in primary care: A population-based cohort study. *PLoS medicine* 2020;17(10):e1003295.

20. Nyante SJ, Black A, Kreimer AR, et al. Pathologic findings following false-positive screening tests for ovarian cancer in the Prostate, Lung, Colorectal and Ovarian (PLCO) cancer screening trial. *Gynecologic Oncology* 2011;120(3):474-79. doi: <https://doi.org/10.1016/j.ygyno.2010.11.015>

21. NHS England. National Schedule of NHS Costs 2021/2022. 2023

22. Oken MM, Hocking WG, Kvale PA, et al. Screening by chest radiograph and lung cancer mortality: the Prostate, Lung, Colorectal, and Ovarian (PLCO) randomized trial. *Jama* 2011;306(17):1865-73.

23. Corley DA, Jensen CD, Quinn VP, et al. Association between time to colonoscopy after a positive fecal test result and risk of colorectal cancer and cancer stage at diagnosis. *Jama* 2017;317(16):1631-41.

24. Broder MS, Ailawadhi S, Beltran H, et al. Estimates of stage-specific preclinical sojourn time across 21 cancer types. *Journal of Clinical Oncology* 2021;39:e18584.

25. Blackford AL, Canto MI, Dbouk M, et al. Pancreatic cancer surveillance and survival of high-risk individuals. *JAMA oncology* 2024;10(8):1087-96.
